# Common variants near the bradykinin receptor B_2_ gene are associated with angioedema induced by angiotensin-converting-enzyme inhibitor treatment - a genome wide association study

**DOI:** 10.1101/2020.12.10.20246967

**Authors:** Jonas Ghouse, Gustav Ahlberg, Laura Andreasen, Karina Banasik, Søren Brunak, Michael Schwinn, Ina Holst Larsen, Oscar Petersen, Erik Sørensen, Henrik Ullum, Eva Rye Rasmussen, Niclas Eriksson, Pär Hallberg, Mia Wadelius, Henning Bundgaard, Morten S. Olesen

## Abstract

**Objective:** Angioedema is a rare, but potentially life-threatening adverse reaction, associated with angiotensin-converting-enzyme inhibitors (ACEi). Identification of potential genetic factors related to this adverse event may help identify at-risk patients.

**Design, Setting and Participants:** A genome-wide association study (GWAS) involving patients of European descent, all taking ACEi was conducted in a discovery cohort (Copenhagen Hospital Biobank), and associations were confirmed in a replication cohort (Swedegene). Cases were defined as persons with an angioedema event and a filled prescription for an ACEi 180 days prior to the event. Controls were defined as persons with continuous treatment with ACEi without any history of angioedema. Odds ratios (ORs) and 95 % confidence intervals (95 % CI) were computed for angioedema risk by logistic mixed model regression analysis. Summary statistics from the discovery and the replication cohorts were analyzed with a fixed effects meta-analysis model.

**Exposure:** Single-nucleotide polymorphisms associated with ACEi-associated angioedema.

**Main outcome:** Hospital record of angioedema.

**Results:** The discovery cohort consisted of 462 cases and 53,391 ACEi-treated controls. The replication cohort consisted of 144 cases and 1,345 ACEi-treated controls. In the discovery cohort, we identified one locus, residing at chromosome 14q32.2, that was associated with angioedema at the genome-wide significance level of *P <*5 × 10^−8^. The lead variant at this locus, rs34485356, is an intergenic variant located 60 kb upstream of *BDKRB2* (OR 1.62; 95 % CI 1.38 to 1.90; *P* = 4.3 × 10^−9^). This variant was validated in our replication cohort with similar direction and effect size (OR 1.60; 95 % CI 1.13 to 2.25; *P* = 7.2 × 10^−3^). We found that carriers of the risk allele had significantly lower systolic (−0.46 mmHg per T allele; 95 % CI −0.83 to −0.10; *P* = 0.013) and diastolic blood pressure (−0.26 mmHg per T allele; 95 % CI −0.46 to −0.05; *P* = 0.013).

**Conclusion:** In this GWAS, involving individuals treated with ACEi, we found that common variants located in close proximity to the bradykinin receptor B_2_ gene were associated with ACEi-related angioedema. *BDKRB2* genotype-directed therapy may aid in improving safety in evidence-based clinical decision-making.

## INTRODUCTION

Angiotensin-converting-enzyme inhibitors (ACEi) are one of the most frequently prescribed drug classes on the market mainly due to its broad indications, including hypertension, heart failure and chronic renal disease. A rare side effect to treatment with this drug class is angioedema, which has been estimated to affect up to 0.7 % of patients treated with ACEi.[1,2] Most patients that experience angioedema have onset within weeks to months of drug initiation; however, the first episode of angioedema can occur after several years of treatment.[3] Although the absolute risk of ACEi–associated angioedema is indeed low, due to the widespread use of ACEi, a comparatively large group of patients are at risk of this condition.[4] In fact, ACEi account for one third of all cases admitted to the emergency room with angioedema.[5] ACEi-associated angioedema most often involves the upper aerodigestive tract but can, in rare cases, affect the bowel.[5] Obstruction of the upper airway occurs in 10 % of cases and can ultimately lead to airway compromise and death.[5,6]

Although the precise mechanism by which ACEi lead to angioedema is not known, accumulation of bradykinin as a result of ACE-inhibition is thought to play a central role.[7] Increased levels of bradykinin cause vasodilation, increased vascular permeability and plasma extravasation into the submucosal tissue, resulting in angioedema.[7,8] Despite this, it remains unclear why angioedema only affects a very small proportion of patients, given that bradykinin levels are reported to increase during ACEi-treatment. This discrepancy has partly been attributed to environmental risk factors that contribute to individual susceptibility to angioedema. Such risk factors include smoking, ethnicity, history of seasonal allergy, previous drug-induced rash, and use of immunosuppressive drugs.[9] Genetic predisposition has also been considered as a risk factor for angioedema. In fact, through early candidate gene studies, variations in the genes *XPNPEP2, MME, ETV6, F5* have been associated with ACEi-associated angioedema.[10–15] Although many of these loci are biologically plausible, only the *XPNPEP2* locus, located on the X chromosome, has successfully been replicated.[11,13,14] In recent years, two genome-wide association studies (GWAS) have been conducted on ACEi-associated angioedema, however, none of the findings reported in these two studies were validated in independent replication cohorts.[12,16]

The increasing availability of genetic and phenotypic data in large biobanks provides an opportunity for investigating the role of genetic variation in rare drug-induced adverse effects. In the present study, we sought to identify genetic risk factors underlying ACEi-associated angioedema.

## METHODS

### Study cohorts

The discovery cohort is part of the “Genetics of cardiovascular disease” – a genome-wide association study on repository samples from Copenhagen Hospital Biobank (CHB), selected based on a history of cardiovascular disease (e.g. ischemic heart disease, hypertension, cardiac arrhythmia, lipid metabolism disorders etc., registered in the Danish National Patient Registry [more information in **Supplementary Methods**]).[17] Patients included in CHB have been informed about their right to refuse the use of their samples for research. Individuals registered in the national opt-out registry were excluded from analysis. The study was approved by The Danish National Ethical Committee (1708829) and the Danish Data Protection Agency (general approval number 2012-58-0004, and local number: RH-2007-30-4129/I-suite 00678). We used Swedegene for replication. Briefly, Swedegene was established in 2008, and consists of individuals who have experienced a serious adverse drug reaction (ADR) and have been reported to the Swedish Medical Products Agency by health care professionals or identified through active surveillance in routine health care.[18]

### Case and control ascertainment

In the discovery cohort, we defined cases as those with a registry diagnosis of angioedema and a redeemed ACEi prescription within 180 days prior to the event. The control group was defined as individuals treated with ACEi for at least two years and no history of angioedema **(S1 Figure)**. In the replication cohort, individuals with angioedema while treated with ACEi were selected as cases. Controls were obtained from the Swedish Twin registry.[19] Unrelated controls were selected if they had at least two consecutive redeemed prescriptions of an ACEi and no history of angioedema. Data on ATC codes to select drugs and ICD-10 codes used to define angioedema are provided in **S1 and S2 Table**, respectively. Information on the registries that have been employed are available in the **Supplementary Methods**.

### Genotyping and imputation

The CHB samples were genotyped using the Global Screening Array by Illumina. Quality control of the CHB data set has been described in detail previously.[20] In brief, samples with non-European genetic ancestry, individuals with sex chromosome aneuploidies or mismatch between inferred sex and sex chromosome count, or outliers for heterozygosity were excluded. The genotyped data were long-range phased using Eagle2[21] and imputed using a reference panel backbone consisting of whole-genome sequence data from 8,429 Danes along with 7,146 samples from Northwestern Europe.[20] Whole genome sequencing, chip-genotyping and the subsequent imputation from which the data for this analysis were generated was performed at deCODE genetics. Details on genotyping, imputation, and association analysis for the replication cohort have been described elsewhere.[16]

### Association analysis

Prior to association testing, additional post-imputation filters included minor allele frequency (MAF) > 1% and imputation score (INFO) > 0.7. After variant-level quality control, ∼12.1 million imputed autosomal variants remained for analysis. We used the Scalable and Accurate Implementation and Generalized mixed model (SAIGE) method[22], which accounts for cryptic population substructure, sample relatedness and controls for inflated Type 1 errors, which can arise with imbalanced case-control ratios. Sex, age and the first 10 principal components (PCs) were used as covariates.

### Replication and meta-analysis

To assess the reproducibility of our findings, the lead variant that reached the genome-wide significant threshold (*P* < 5 × 10^−8^) was tested in the Swedegene GWAS data on ACEi-associated angioedema.[16] Significance threshold for replication was set at *P <* 0.05. To maximize the statistical power to detect associated genetic variants of small effect, we further conducted a meta-analysis of the two interrogated cohorts using summary statistics from the discovery and replication data. Prior to meta-analysis, individual GWAS results were assessed using the R package EasyQC (v9.2), and duplicates, monomorphic SNPs and copy-number variations were excluded.[23] Only SNPs with MAF > 1% and INFO > 0.7 were moved forward for meta-analysis. The absence of population stratification was controlled for based on the genomic inflation factor (λ < 1.10 for both cohorts). More than ∼8.8 million high-quality SNPs were available for meta-analysis which was performed with METAL using the fixed-effect inverse variance weighted method with *z* scores weighted by sample size.[24]

### Functional annotation

FUMA (version 1.3.6) [25] was used for functional annotation of the GWAS meta-analysis results. Before defining potential genomic risk loci, FUMA defines independent significant variants as variants that are genome-wide significant (*P*□<□5□×□10^−8^) and are independent (*r*^2^□< □0.6). Within this pool of independent SNPs, lead SNPs are defined as those in linkage disequilibrium (LD) at *r*^2^□<□0.1. Candidate SNPs were defined as SNPs in the risk locus with a GWAS P-value of <10^−5^ and LD r^2^ >0.6 with one of the independent significant SNPs. Genotypes from the 1000G phase 3 were used as reference data to infer LD. Genetic risk loci were defined by combining lead SNPs within a 250□kb window and all SNPs in LD of at least *r*^2^□=□0.6 with one of the independent SNPs. SNPs were annotated with Annotate Variation (ANNOVAR)[26], Combined Annotation Dependent Depletion (CADD) scores[27], RegulomeDB[28] scores, and chromatin state[29,30]. More details on these databases are available in the **Supplementary Methods**.

### Gene mapping

Loci obtained from the GWAS meta-analysis were mapped to genes using four different approaches:

1. *Positional mapping*: Maps SNPs to genes based on physical distance (± 100 kb) from known protein-coding genes in the human reference assembly (GRCh37/hg19).
2. *Chromatin interaction mapping*: Maps SNPs to genes, when there is a three-dimensional DNA-DNA interaction between the SNP region and another gene region. We used Hi-C data of 23 different cell and tissue types embedded in the FUMA platform.[31] We used a false discovery rate of <1 × 10^−6^ to define significant interactions.
3. *Expression quantitative trait loci (eQTL) mapping*: Maps SNPs to genes with which the SNPs show a significant eQTL association (i.e. allelic variation at the SNP is associated with the expression level of the gene in question). We used information from 30 tissue types from the GTEx v8 data repository, which is based on cis-eQTLs that can map SNPs up to 1 megabase apart. We used a false discovery rate of < 0.05 to define significant eQTL associations.
4. *Colocalization analysis:* To prioritize among candidate genes identified through the listed gene mapping strategies, we estimated the posterior probability for a common causal variant underlying association with gene expression and ACEi-associated angioedema, by conducting pairwise Bayesian colocalization analysis, implemented in the *coloc* R package v.3.1.[32] We used publicly available genome-wide eQTL data from the GTEx v8 portal. Tissues highlighted with significant eQTL-associations with our risk locus were prioritized. We tested all genes with significant *cis*-eQTL association by analyzing all variants within a ± 250 kb window around our risk locus using eQTL summary data, and ACEi-associated angioedema summary data from the present study. Default priors were used. A posterior probability of ≥ 0.7 was set as threshold to support a causal role for the gene as a mediator of angioedema risk.

### Gene-region-based testing

A gene-region-based association analysis (GWGAS) was performed using MAGMA[33], also embedded in the FUMA platform.[25] Herein, variants were assigned to protein-coding genes (n=19,294) if they were located ± 50 kb from the gene body. The resulting *P*-values were combined into a gene test-statistic using the SNP-wise mean model. We accounted for multiple testing, by defining threshold for significance at *P* < 2.6 × 10^−6^ (0.05/19,294).

### Heritability

Genome-wide SNP heritability (h^2^_g_) was calculated using restricted maximum likelihood, implemented in BOLT-REML.[34] We used a genetic relationship matrix that included 331,583 genotyped model SNPs from our discovery cohort, with a MAF above 1 % and genotyping rate across subjects > 99 %, and the pre-computed LD scores based on 1000G European descent data. We adjusted for sex, age and 10 PCs.

### Association between lead variant and blood pressure in individuals treated with ACE-inhibitors

To evaluate the aggregate effects of treatment with ACEi and the risk allele, we used data on unrelated individuals of European ancestry with automated blood pressure readings and filled prescriptions for ACE-inhibitors (**S1 Table**) in the UK Biobank (UKB). Samples were excluded based on centralized sample quality control performed by UKB as follows: if inferred sex did not match reported sex, kinship was not inferred, putative sex chromosome aneuploidy, excessive heterozygosity or missingness.[35] Related individuals were defined as individuals with *KING* coefficient > 0.0884, indicating 2^nd^ degree or closer relatedness. European ancestry was determined by self-reported ancestry of British, Irish, or other white. We excluded population outliers, by defining means and standard deviations (SDs) on PCs 1-5 of the European ancestry population. Individuals with a sum total of more than 6 SDs in PCs 1-5 were excluded. We used linear regression to evaluate the effect of rs34485356 on systolic and diastolic blood pressure per T allele increase, respectively, and adjusted the model for age, sex, 10 PCs, assessment center and genotyping array.

### Mendelian randomization

Multiple factors have shown either protective or risk increasing effects associated with ACEi-induced angioedema in observational studies.[36] However, only smoking, seasonal allergy, coronary artery disease (CAD) and type 2 diabetes (T2D) have consistently been reported as plausible risk or protective factors.[37] To evaluate the causal effects of these traits on angioedema risk, we used Mendelian randomization (MR). All MR analyses were performed using the TwoSampleMR package.[38] The inverse variance weighted method was used as the primary method,[39] and the weighted median estimation and MR-Egger were used as sensitivity analyses.[40] We also used the MR Pleiotropy RESidual Sum and Outlier (MR-PRESSO) method to assess whether any detectable effect was mediated through outliers.[41] To select independent SNPs for each exposure, GWAS significant SNPs (*P* < 5 × 10^−8^) for each risk factor was pruned using default parameters (*r*^*2*^ < 0.001 and an LD window of 10,000 kb). SNPs that were unavailable in our angioedema summary data were replaced with a suitable proxy (*r*^*2*^ ≥ 0.8), where available. A *P <* 0.05 was considered as suggestive evidence for association. Summary statistics and SNPs that were used as instruments for each exposure are summarized in **S3-S7 Table**. A detailed description of the MR methods employed in this study is provided in the **Supplementary Methods**.

## RESULTS

### Clinical characteristics

A total of 462 individuals from the CHB had experienced angioedema while on treatment with ACEi (54.1 % men; median [IQR] age 77.3 [70.6, 84.5]; **Table 1**). A total of 53,391 ACEi treated individuals without any history of angioedema constituted the control group for our discovery cohort (57.7 % men; median [IQR] age 75.9 [67.7-84.3]). We found that cases were more likely older and had higher proportion of prevalent hypertension and myocardial infarction, compared with controls (**Table 1**). The most commonly prescribed drug among cases and controls was enalapril, followed by ramipril. Cases were more often prescribed combination therapy with diuretics than controls. The replication cohort consisted of 144 individuals with a history of angioedema while treated with ACEi, and 1,345 ACEi-treated controls without any history of angioedema. Characteristics of the replication cohort is provided in **S8 Table**.

**Table 1.** Participant characteristics of ACE inhibitor-associated angioedema cases and ACE inhibitor-treated controls in the Copenhagen Hospital Biobank on January 1, 2019.

| **Clinical characteristics** | **Cases**<br>***n*=462** | **Controls**<br>***n*=53,391** |
| --- | --- | --- |
| Age, median (IQR) | 77.3 (70.6, 84.5) | 75.9 (67.7, 84.3) |
| Male sex, *N* (%) | 250 (54.1) | 30,787 (57.7) |
| *Comorbidity, N (%)* |  |  |
| Hypertension | 411 (89.0) | 41,762 (78.2) |
| Diabetes | 144 (31.2) | 14,654 (27.4) |
| Heart failure | 125 (27.1) | 16,370 (30.7) |
| Myocardial infarction | 79 (17.1) | 11,876 (22.2) |
| Chronic renal disease | 78 (16.9) | 6,485 (12.1) |
| *Type of ACE-inhibitor, N (%)* |  |  |
| Plain therapy |  |  |
| C09AA01, Captopril | 2 (0.4) | 507 (0.9) |
| C09AA02, Enalapril | 214 (46.3) | 27,726 (51.9) |
| C09AA03, Lisinopril | 8 (1.7) | 1,523 (2.9) |
| C09AA04, Perindopril | 9 (1.9) | 2,589 (4.8) |
| C09AA05, Ramipril | 73 (15.8) | 11,379 (21.3) |
| C09AA06, Quinapril | 0 (0.0) | 60 (0.1) |
| C09AA07, Benazepril | 0 (0.0) | 24 (0.0) |
| C09AA09, Fosinopril | 0 (0.0) | 67 (0.1) |
| C09AA10, Trandolapril | 37 (8.0) | 4,282 (8.0) |
| Combination therapy |  |  |
| C09BA01, Captopril and diuretics | 0 (0.0) | 20 (0.0) |
| C09BA02, Enalapril and diuretics | 82 (17.7) | 3,154 (5.9) |
| C09BA03, Lisinopril and diuretics | 14 (3.0) | 493 (0.9) |
| C09BA04, Perindopril and diuretics | 1 (0.2) | 414 (0.8) |
| C09BA05, Ramipril and diuretics | 22 (4.8) | 1,141 (2.1) |
| C09BB02, Enalapril and lercanidipine | 0 (0.0) | 6 (0.0) |
| C09BB04, Perindopril and amlodipine | 0 (0.0) | 6 (0.0) |

ICD-10 codes to define comorbidities are provided in **S13 Table**. ACE, angiotensin-converting-enzyme; IQR, interquartile range; N, number of individuals.

### Genome-wide association analysis of ACE-inhibitor-associated angioedema

We identified one novel locus, residing in chromosome 14q32.2, that was associated with angioedema at the genome-wide significance level of *P <*5 × 10^−8^ (**S2 Figure**, λ_GC_ = 0.99). The lead variant at this locus, rs34485356, is an intergenic variant located 60 kb upstream of *BDKRB2* (**Table 2**; OR 1.62; 95 % CI 1.38-1.90; *P* = 4.3 × 10^−9^). This variant replicated in our replication cohort with similar effect size and direction of effect (**Table 2**; OR 1.60; 95 % CI 1.13-2.25; *P* = 7.2 × 10^−3^). **Figure 1A** and **Table 2** display the main results from the meta-analysis. Meta-analysis of the two GWA studies did not reveal additional genome-wide significant loci. Nor did variants previously linked with ACEi-associated angioedema replicate in this study **(Supplementary Methods** and **S9 Table)**. SNP-based heritability estimates calculated using BOLT-REML was *h*^*2*^*=* 21.7 % (SE 0.097). A QQ plot for each GWAS (discovery and replication) is displayed in **S3 Figure** and a regional plot for the genome-wide significant locus is provided in **S4 Figure**.

**Table 2.**
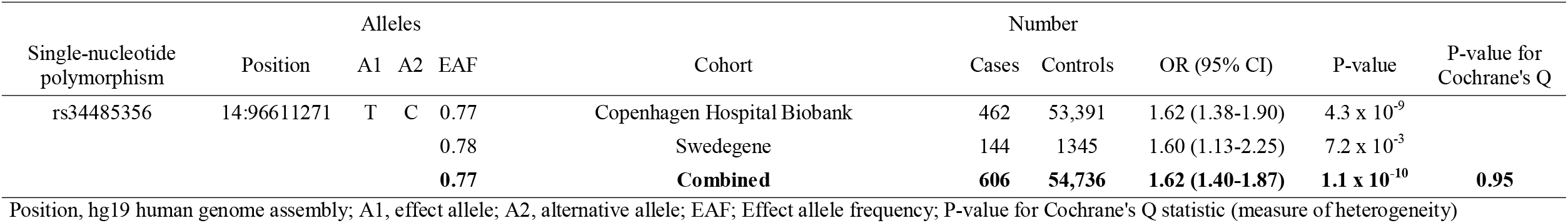
Association results with ACE-inhibitor associated angioedema in discovery (Copenhagen Hospital Biobank) and replication (Swedegene) and combined analysis

**Figure 1.**
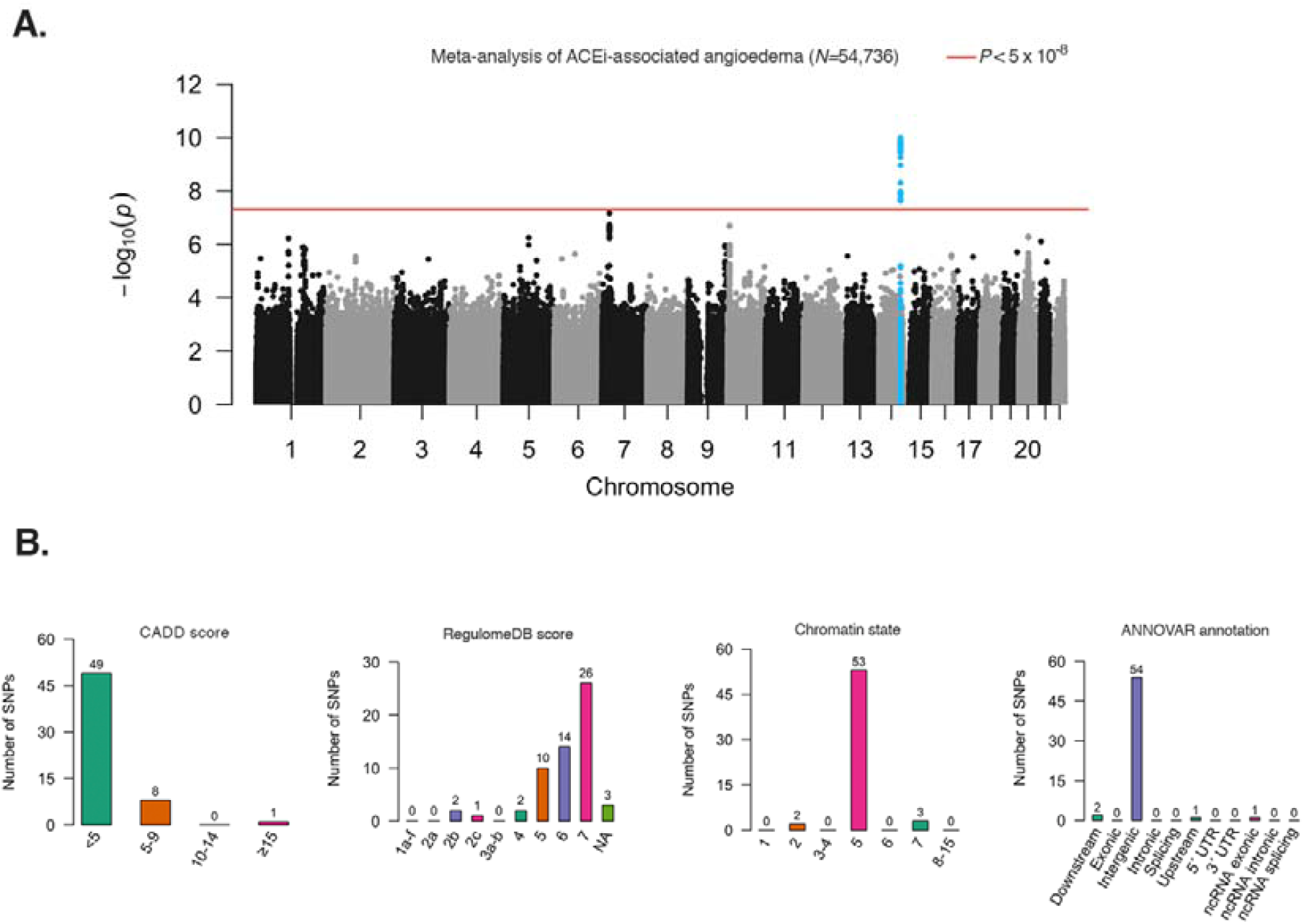
SNP-based results from the genome-wide association study (GWAS) meta-analysis on ACE-inhibitor associated angioedema in 54,736 individuals. **A**. Manhattan plot of the GWAS meta-analysis of ACE-inhibitor associated angioedema, showing the negative log10-transformed *P*-value for each SNP. *P*-values were obtained from a logistic regression mixed model. **B**. Distribution of CADD scores, RegulomeDB categories, chromatin state and functional consequences of all annotated SNPs in linkage disequilibrium (r^2^ ≥ 0.6) with the lead GWAS SNPs.

### Functional annotation of associated variants and gene mapping

We performed functional annotation using FUMA[25] on our meta-analyzed data. We identified a total of 58 candidate SNPs. The majority of these SNPs were most abundant in intergenic regions (n=54, 93.0 %, **Figure 1C**), and most overlapped with open chromatin state annotations (**Figure 1C**), suggesting that they may affect gene regulation. One of the candidate SNPs, the proxy variant rs72704813 (*P =* 1.6 × 10^−10^, r^2^ = 1.0 with the lead SNP; **S10 Table**), had the lowest RegulomeDB score (2b; likely to affect binding; **S10 Table**), indicating that impaired binding of transcription factors may be involved. This proxy also had the highest CADD score (18.11; **S10 Table**), indicating that variation at this locus is associated with deleterious effect.

To gain insights into the functional consequences of our risk locus, we mapped variants to genes by proximity, eQTL mapping, and chromatin interaction (see **Methods**). Three genes, *C14orf132, RP11-404P21*.*8*, and *BDKRB2* (**S11 Table**) were naively mapped based on a ±100 kb distance from our risk locus. eQTL gene mapping matched cis-eQTL variants to two nearby genes (*BDKRB1* and *BDKRB2;* **Figure 2A** and **S11 Table**). Chromatin interaction mapping annotated five genes (*RP11-1070N10-3, RP11-404P21*.*8, BDKRB2, GLRX5*, and *TCL1A)*, based on DNA-DNA interactions between our risk locus and nearby or distant genes (**Figure 2A, S12 Table and S13 Table)**. In addition to linking individually associated variants to genes, we also conducted a GWGAS using MAGMA (see **Methods**). In our GWGAS, apart from the long non-coding RNA *RP11-404P21*.*8* (*P* = 4.5 × 10^−7^), the *BDKRB2* (*P* = 1.5 × 10^−6^) was the only gene that passed the multiple-testing threshold (**Figure 2B** and **S14 Table**). To add further support that our risk locus mediates its effect through *BDKRB2*, we estimated the posterior probability for a common causal variant underlying the association between our risk locus and angioedema, by conducting pairwise Bayesian colocalization analysis. We found evidence for colocalization for both *BDKRB1* and *BDKRB2* in brain tissue **(S12 Table)**, indicating that our risk locus affects gene expression. The *BDKRB2* gene was the only gene that was implicated by positional mapping, eQTL-associations, chromatin interactions, GWGAS *and* colocalization analysis, indicating that our risk locus is most likely associated with this gene.

**Figure 2.**
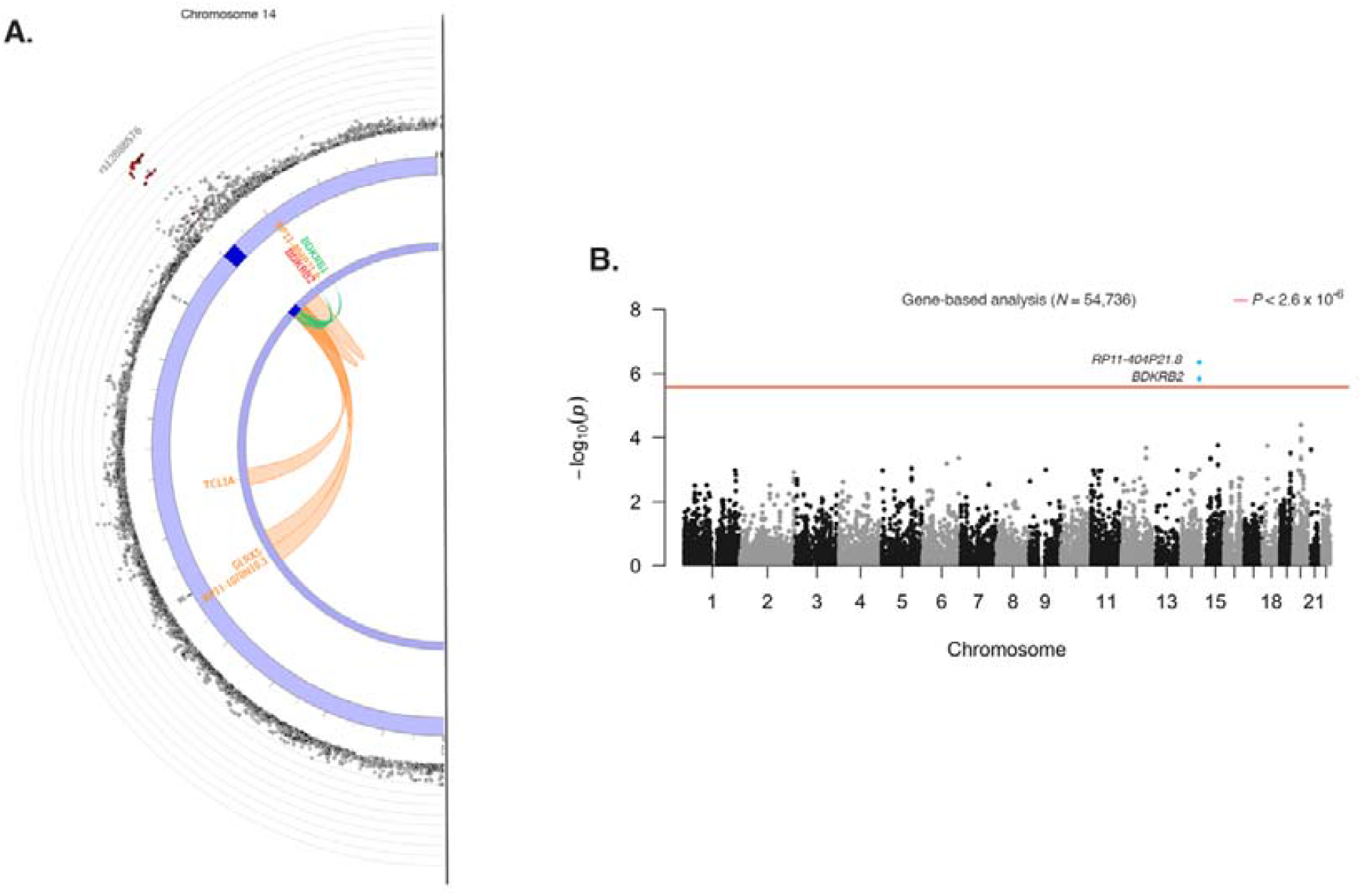
Gene-based analysis of ACE-inhibitor associated angioedema in 54,736 individuals. **A**. Zoomed-in circo plots showing the genes implicated by the genomic risk locus on chromosome 14, with the genomic risk locus indicated with a blue shaded area, eQTL associations in green, and chromatin interactions in orange. Genes mapped by both eQTL and chromatin interactions are red. The outer layer shows a Manhattan plot containing the negative log10-transformed *P-*value of each SNP in the GWAS analysis of ACE-inhibitor associated angioedema. **B**. Genome-wide gene-based analysis (GWGAS) of genes (*N*=19,294) that were tested for association with ACE-inhibitor associated angioedema in MAGMA. The y-axis shows the negative log10-transformed two-sided *P*-value of the gene-based test, and the x-axis shows the starting position on the chromosome. Gene-based two-sided *P*-values were calculated with MAGMA. The red line indicates the Bonferroni corrected threshold for genome-wide significance (2.6 × 10^−6^ [0.05/19,294]). Genes that passed the significance threshold are highlighted.

### Effect of rs34485356 on blood pressure

To evaluate the aggregate effects of ACE-inhibition and increased expression of the *BDKRB2* gene, we specifically tested whether a subgroup of ACEi-treated persons, with drug-induced increased levels of bradykinin, who also carried the risk allele, would differ in blood pressure, compared with people without the risk allele. In total, we identified 34,873 unrelated individuals on ACEi-therapy with available systolic and diastolic blood pressure measurements in the UKB. We found that carriers of the risk allele had significantly lower systolic (−0.46 mmHg per T allele; 95 % CI −0.83, −0.10; *P* = 0.013) and diastolic (−0.26 mmHg per T allele; 95 % CI −0.46, −0.05; *P* = 0.013) blood pressure compared with non-carriers.

### Causal influence of risk factors on ACEi-associated angioedema

We performed MR analyses to evaluate the potential causal role of four pre-defined cardiometabolic traits on ACEi-associated angioedema risk (selected based on previous observational evidence linking such traits to angioedema risk). We found an association between genetic liability to T2D and lower risk of angioedema (**Figure 3**, IVW; OR 0.82 [95% CI 0.67, 0.99]). Sensitivity analyses showed estimates that were consistent both in magnitude and direction of effect. We found no indications of a causal relationship between genetic liability to smoking, allergy and CAD and risk of ACEi-associated angioedema **(Figure 3)**.

**Figure 3.**
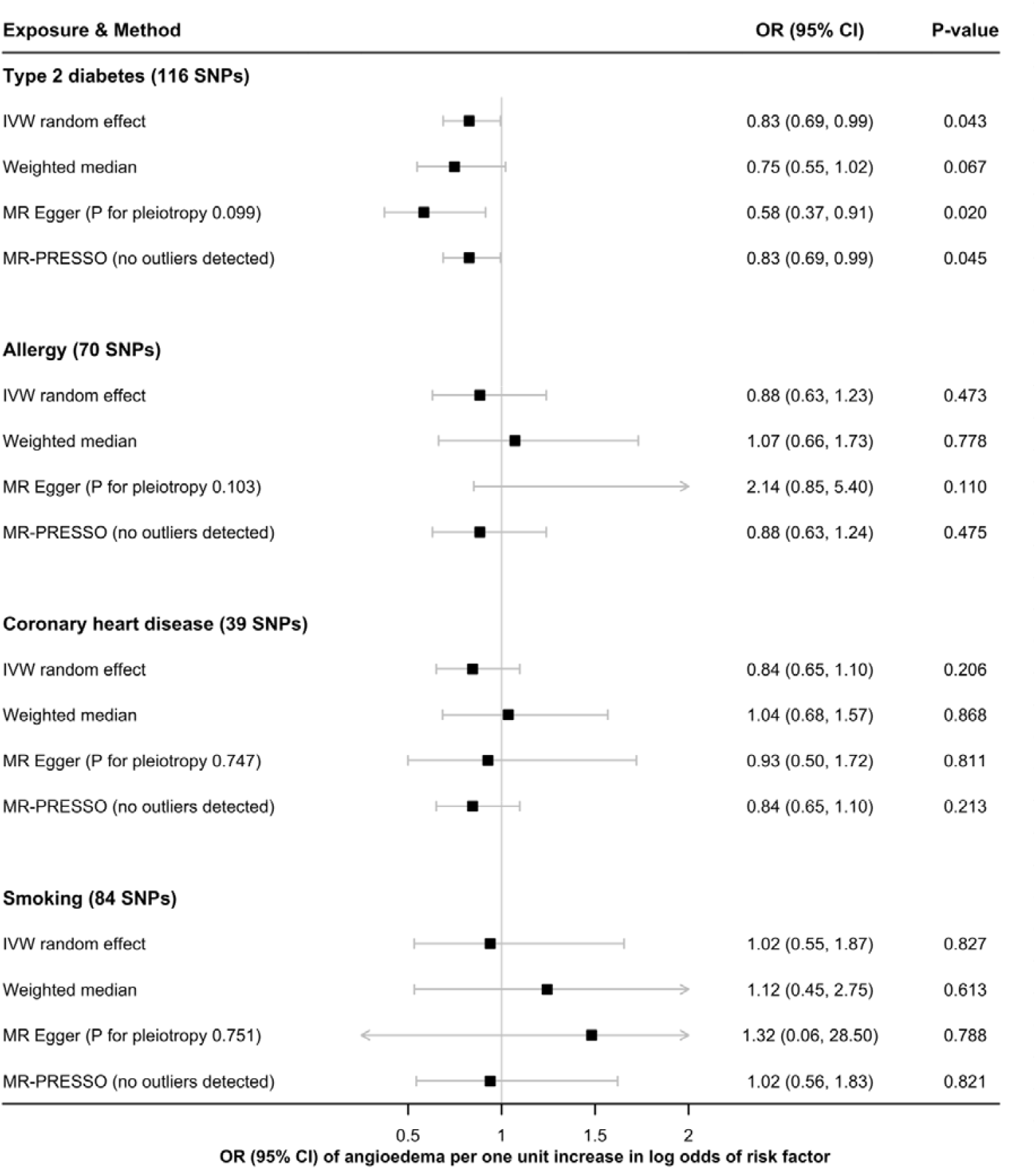
Association between four cardiometabolic traits and ACE-inhibitor-associated angioedema risk using Mendelian randomization. Squares represent odds ratios and error bars indicate the 95 % confidence intervals.

## DISCUSSION

By leveraging the largest genome-wide data to date on this ACEi-associated adverse event, we identified the first replicated genome-wide significant association between genetic variation at the bradykinin receptor B_2_ locus and ACEi-associated angioedema.

The primary target of ACEi is the renin-angiotensin-aldosterone system, which results in decreased production of angiotensin II and leads to decreased aldosterone secretion, decreased vasopressin activity, and the favorable antihypertensive and cardiovascular effects that are seen in this drug class.[42] The mechanism of ACEi-associated angioedema is thought to be related to its role in the kallikrein-kinin system, an antagonistic counterpart to the renin-angiotensin-aldosterone system. Kallikrein is a protease that converts high-molecular-weight kininogens into kinins, primarily bradykinin. Bradykinin is a potent vasodilator because it mediates the release of nitric oxide, substance P, prostacyclin, and endothelium-derived hyperpolarizing factor via binding to bradykinin receptor B_2_.[42] These agents facilitate increased vascular permeability, which ultimately results in the localized inflammation and edema that characterize angioedema.[42] Normally, this response is controlled by the action of angiotensin-converting enzyme (or kininase II), which degrades bradykinin. However, despite decreased metabolism of bradykinin during ACE-inhibition, only a fraction of patients on ACEi develop angioedema, indicating that other important factors may be involved. In this study, we found an association between a locus at chromosome 14q32.2 and ACEi-associated angioedema. We provide several lines of evidence (e.g. eQTL association and colocalization analyses) that support that *BDKRB2*, located 60 kb downstream of our lead variant, is the likely causal gene at this locus. A simple model to explain these observations would be that the *BDKRB2-*variant leads to increased expression of the bradykinin B_2_ receptor or increased sensitivity to bradykinin, ultimately leading to increased susceptibility for vasodilation and fluid extravasation. The observed parallel effect of significantly lower systolic and diastolic blood pressure in people carrying the risk allele *and* treated with ACEi, also suggests that the risk locus not only predisposes to localized vasodilation when treated with ACEi, but also to systemic effects on blood pressure.

Several other genes, including *XPNPEP2, MME, F5, ETV6, KCNMA1*, have been implicated in ACEi-associated angioedema, with biologically plausible roles in angioedema pathology.[10–16] However, the majority of genes have been linked with angioedema through candidate gene methods, and only two studies to date, have used a GWAS approach.[12,16] None of the studies that have investigated the role of common variants in autosomal genes have been successfully replicated in independent cohorts. Nor did any of the variants associate with ACEi-associated angioedema in this study. Although we found one genetic locus associated with angioedema risk, given the relatively high heritability estimate, potentially additional genetic loci remain to be identified.

Apart from genetic polymorphisms, other environmental risk factors, such as smoking, concomitant use of immunosuppressants, ethnicity, seasonal allergy, history of drug rash, prevalent cardiopulmonary disease and female sex, may also be of importance.[9,37] In this study, we evaluated the causal relationship between four predefined risk factors and ACEi-associated angioedema, using an MR approach. These risk factors were selected based on the strength of the association and availability of summary statistics to evaluate the relationship. We found evidence of a causal relationship between genetic liability to T2D and lower risk of ACEi-associated angioedema. Studies have reported that exposure to high glucose levels increases the expression of dipeptidyl peptidase 4 (DPP-4), an enzyme that plays a central role in the breakdown of both incretins and bradykinin.[43] Conversely, co-administration of DPP-4 inhibitors with ACEi or angiotensin receptor blockers have been associated with an exacerbated risk of angioedema, most likely due to reduced clearance of bradykinin.[44] We found limited evidence of a causal relationship between allergy, smoking or CAD and angioedema risk. However, an additive effect of these risk factors with the genotype cannot be excluded, and warrants further investigations.

Our study has implications on clinical care. Seventy-eight percent of the European population and 81 % of Africans are carriers of the risk allele, respectively, whereas Asians almost exclusively carry the risk allele (allele frequency 90-100 %).[45] Interestingly, ACEi-associated cough, which is believed to be due to a similar bradykinin-mediated pathway,[46] is 2.5 times more common in Asians than in Caucasians,[47] and withdrawal rates of ACEi exceed 30 %.[48] Thus, adverse events due to this variant may be particularly important in Asian populations. However, the strength of effect of the *BDKRB2* variant on angioedema risk may depend on the aggregate effects of genetics and other risk factors, such as environmental exposures, which may vary among ethnic populations. Our cohort of European descent was not sufficient to examine ethnicity-specific differences on ACEi-associated angioedema risk. Additional studies in diverse populations are therefore warranted.

Since more than 40 million patients are treated with ACEi worldwide, which could potentially result in several hundred fatalities attributed to angioedema, better predictors are needed to aid in clinical decision making. Genotype-directed decision making may prove useful in assisting clinicians to choose the most effective antihypertensive drug, with the lowest risk for severe adverse events for a given individual. Given the frequency of the risk allele, genotype-guided therapy based solely on the risk variant identified in this study is not feasible. However, with the discovery of additional credible genetic determinants of ACEi-associated angioedema, a panel of genetic variants may be used along with clinical data to guide antihypertensive therapy, which does not exert its effects through ACE-inhibition.

In conclusion, in this GWAS, we show that common variants near *BDKRB2* are associated with increased risk of angioedema in ACEi-treated individuals. Additional studies will be necessary to determine whether *BDKRB2* genotype directed therapy can aid in improved safety in evidence-based clinical decision making.

## Data Availability

Data will be made available upon request to the corresponding author.

## Notes

### Competing Interest Statement

The authors have declared no competing interest.

### Funding Statement

J.G is the guarantor of this study. This work was supported by The John and Birthe Meyer Foundation, The Research Foundation at Rigshospitalet, Villadsen Family Foundation, The Arvid Nilsson Foundation, The Hallas-Moeller Emerging Investigator Novo Nordisk (NNF17OC0031204), Novo Nordisk Foundation (grants NNF17OC0027594 and NNF14CC0001), The Swedish Research Council (Medicine 521 2011 2440, 521 2014 3370 and 2018 03307), Swedish Heart and Lung Foundation (20120557, 20140291 and 20170711), Medical Products Agency, Selanders Foundation, Thureus Foundation and Clinical Research Support (Avtal om Laekarutbildning och Forskning) at Uppsala University. The Swedish Twin Registry is managed by Karolinska Institutet, and receives funding through the Swedish Research Council under grant no. 2017-00641.

### Author Declarations

The study was approved by The Danish National Ethical Committee (1708829) and the Danish Data Protection Agency (general approval number 2012-58-0004, and local number: RH-2007-30-4129/I-suite 00678).

