## Supplementary Material for "Common variants near the bradykinin receptor B_2_ gene are associated with angioedema induced by angiotensin-converting-enzyme inhibitor treatment - a genome wide association study"

By Ghouse J *et al*.

**TABLE OF CONTENT**

**SUPPLEMENTARY METHODS**

#### *Data resources*

In Denmark, all citizens are allocated a unique civil registration number. This enables linkage to nationwide administrative registries on an individual basis. The National Population Registry contains information on sex, date of birth, date of emigration and date of death on all Danish citizens.^1^ The Danish National Patient Registry (NPR), established in 1978, holds nationwide records of all hospital, out-patient clinic, and emergency room admissions. Upon discharge, each hospitalization or visit at an out-patient clinic or emergency room is registered with a primary discharge diagnosis (and if applicable, secondary diagnoses) classified according to the International Classification of Disease (ICD-10 from 1994). The Danish National Prescription Registry (DNPR) was established in 1994, and holds detailed information on dispensing date, strength, quantity, and drug type (using the Anatomical Therapeutic Chemical System), on all claimed drug prescriptions dispensed from pharmacies in Denmark.^2^ Using these registries, we obtained information on comorbidity and concomitant drug therapy.

#### *Functional annotation databases*

ANNOVAR categorizes SNPs based on their genic position (e.g. intron, exon, intergenic) and function^3^. CADD scores predict how deleterious the effect of an SNP is likely to be for a protein structure/function, with higher scores representing higher deleteriousness (CADD scores >12.37 suggest that a variant is deleterious)^4^. It integrates information from 63 distinct functional annotations into a single quantitative score, ranging from 1 to 99, based on variant rank relative to all 8.6 billion possible single nucleotide variants of the human reference genome (GRCh37). The RegulomeDB score is a categorical score based on information from expression quantitative trait loci (eQTL) and chromatin marks, which ranges from 1a to 7, with lower scores indicating an increased likelihood of having a regulatory function^5^. The chromatin state represents the accessibility of genomic regions. It is categorized into 15 states predicted by a hidden Markov model based on 5 chromatin marks for 127 epigenomes in the Roadmap Epigenomics Project.^6,7^ A lower state indicates higher accessibility, with states 1 to 7 generally denoted as open state.

#### *Mendelian randomization analyses*

We used four complementary Mendelian randomization methods with distinct strengths and assumptions, to analyze the relationship between genetically-predicted type 2 diabetes, allergy, coronary heart disease and smoking and ACE-inhibitor associated angioedema. The inverse-variance weighted method (under a multiplicative random-effects model) provides the highest precision but assumes that all SNPs are valid instrumental variables.^8^ The weighted median method provides less precise estimates than the inverse-variance weighted method but gives reliable estimates if at least half of the weight in the analysis comes from valid instrumental variables.^8^ We also used the MR-Egger method, which has low precision but can detect and correct for pleiotropy and provide a causal estimate even if all genetic variants have pleiotropic effects on the outcome.^8^ And lastly, the MR-PRESSO (Pleiotropy RESidual Sum and Outlier) method, which can identify and correct for horizontal pleiotropy through removal of outliers.^9^ The analyses were carried out using the MRPRESSO and TwoSampleMR packages^10^.

#### *Phenome-wide association study (PheWAS) analysis*

We queried the UKB GWAS results using the University of Michigan PheWeb web interface (http://pheweb.sph.umich.edu/SAIGE-UKB/). The UKB PheWeb interface comprises results from a SAIGE^19^ genetic analysis of 1,403 ICD-based traits of 408,961 UKB participants of European ancestry. The level of significance was set at *P =* 0.05/1,403 = 3.6 × 10^−5^.

#### *Reappraisal of previously ACEi-associated angioedema variants*

To test whether previously reported ACEi-associated angioedema variants replicate in the present data, we queried our meta-analysis summary data for a predefined list of autosomal SNPs reported in a recent study by Ali *et al.*^43^

### **SUPPLEMENTARY RESULTS**

#### *Phenome-wide association analysis*

We investigated associations between the lead SNP and 1,403 traits in the UKB that may provide additional insight into the aetiology of ACEi-associated angioedema, however none of the associations reached significance beyond the threshold for multiple testing.

### **SUPPLEMENTARY FIGURES**

#### **S1 Figure.** Case and control ascertainment in the discovery cohort (Copenhagen Hospital Biobank).

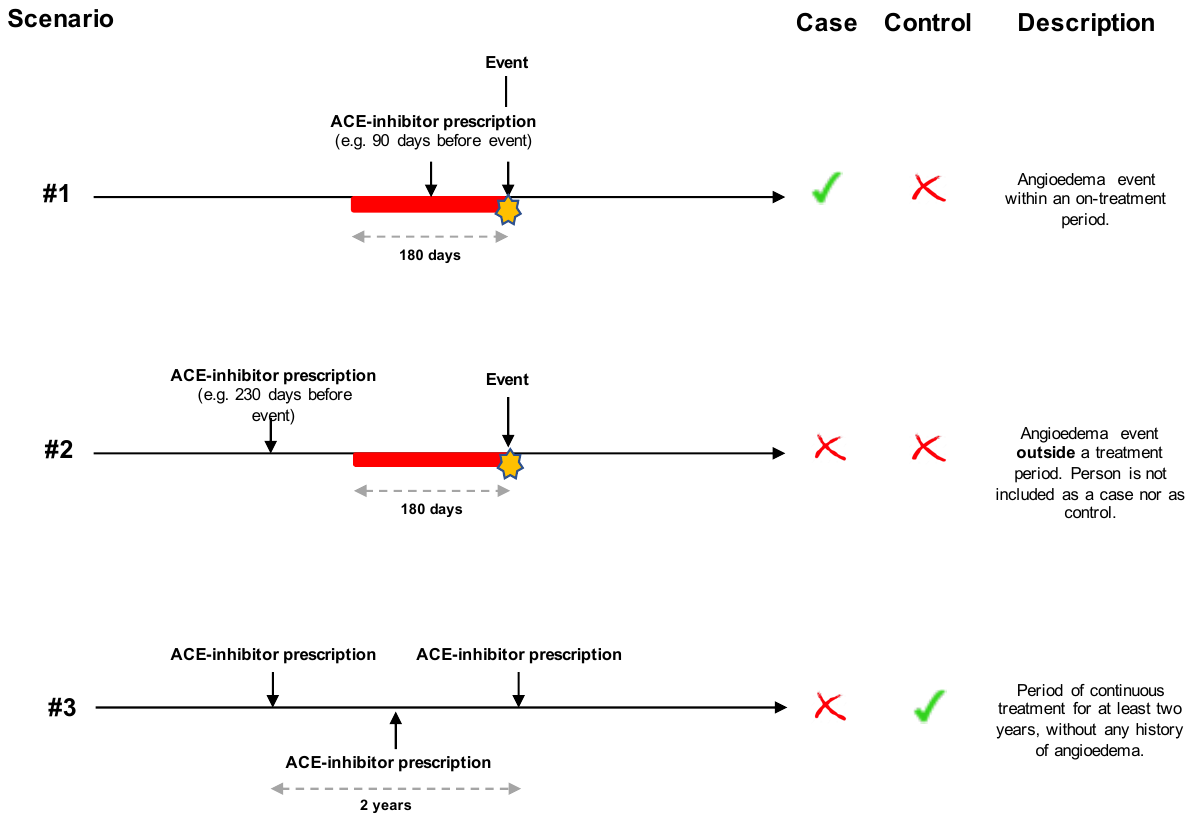

**S2 Figure.** Manhattan plot of the discovery GWAS of ACE-inhibitor associated angioedema, showing the negative log10-transformed P-value for each SNP. P-values were obtained from a logistic regression mixed model.

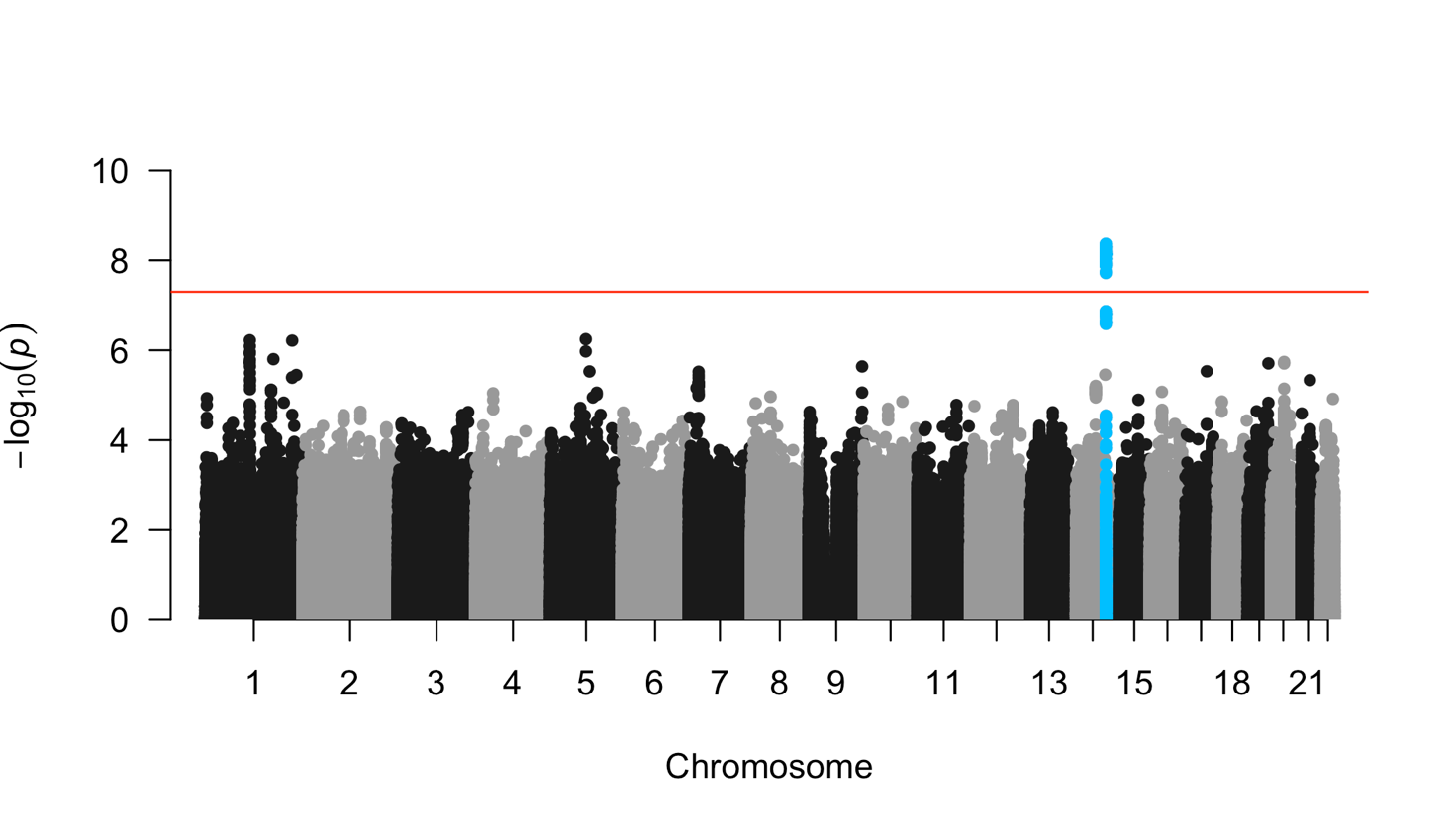

**S3 Figure.** Quantile-quantile plot of A) Discovery GWAS and B) Replication GWAS.
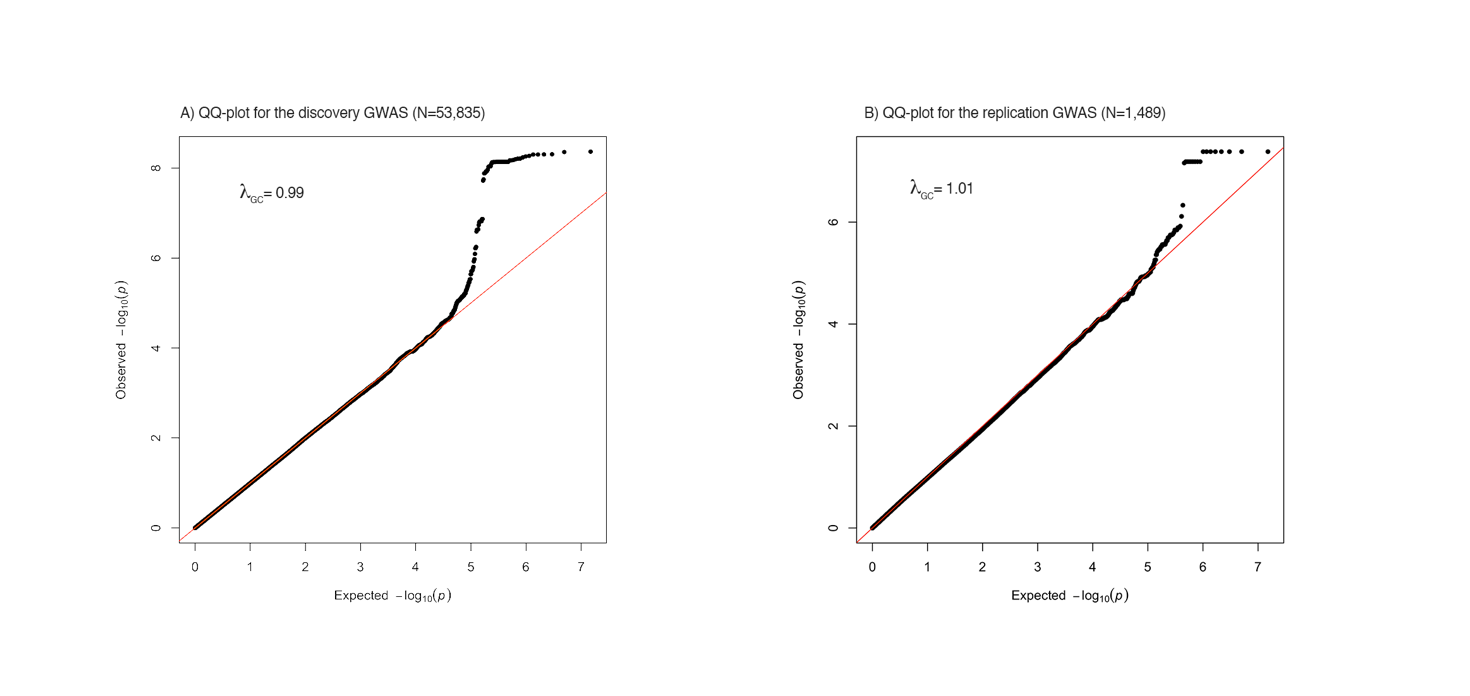

**S4 Figure.** Regional association plot. The x-axis represents a 0.2 Mb region, 100kb on either side of the sentinel variant (rs12888576; purple diamond) and the y-axis shows −log10 P values for individual SNPs. Pairwise LD (r2) with the sentinel variant is based on 1000 Genomes phase 3 v5 European reference samples and is described using the color scale given. The bottom panel show genes located within the region.

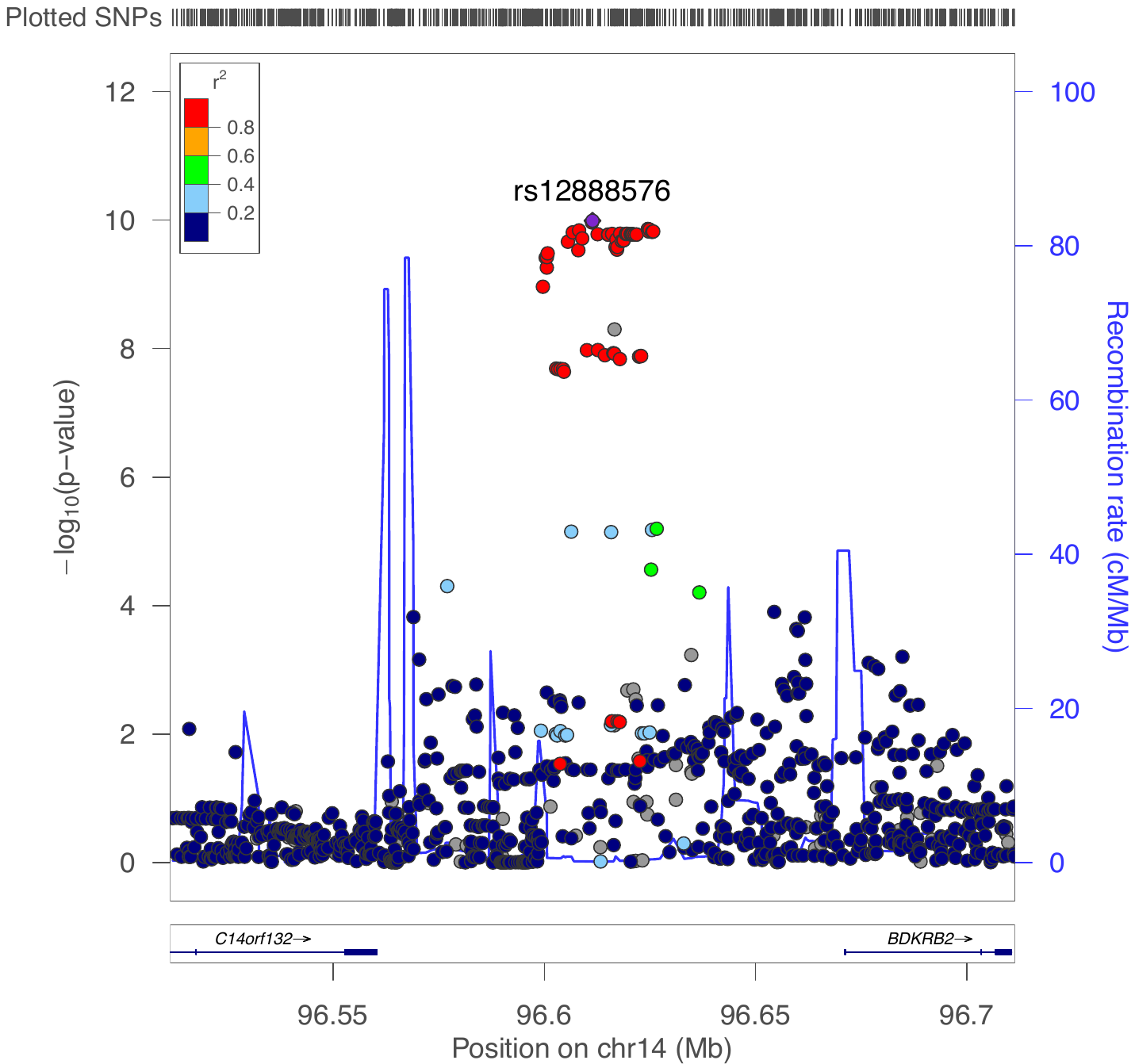

### **SUPPLEMENTARY TABLES**

| **S1 Table.** Identification of drugs from the prescription databases | | |
| --- | --- | --- |
|  | **Database** | **Drug code** |
| ACE-inhibitors | Danish National Prescription Registry (ATC codes) | C09A, C09B |
|  | United Kingdom (UK) Primary Care Prescription data (coded using British National Formulary codes) | 205051 |

| **S2 Table.** Definition of cases from ICD-codes. | | |
| --- | --- | --- |
| **Cohort** | **Registry** | **ICD code** |
| Copenhagen Hospital Biobank | Danish National Patient Registry | **ICD-10:** T783 (Angioneurotic edema), J384 (Laryngeal edema), J392A (Nasopharyngeal edema), J392D (Pharyngeal edema) |
| SwedeGene | Swedish National Patient Registry | **ICD-9:** 9951 (Angioneurotic edema), 4786 (Laryngeal edema); **ICD-10:** T783 (Angioneurotic edema), J384 (Laryngeal edema) |

| **S3 Table.** Information of included studies and consortia in the Mendelian randomization analysis. | | | |
| --- | --- | --- | --- |
| **Exposure** | **Consortium or cohort study** | **Participants** | **Web source if publicly available** |
| Type 2 diabetes mellitus | Meta-analysis of type 2 diabetes, including data from DIAGRAM, GERA, and UK Biobank. | 62,892 cases with type 2 diabetes, and 596,424 controls, all of European ancestry. | <http://cnsgenomics.com/data.html> |
| Coronary heart disease | CARDIoGRAMplusC4D | 60,801 CAD cases and 123,504 controls, mainly of European ancestry. | <http://www.cardiogramplusc4d.org/data-downloads/> |
| Allergy | Meta-analysis of a broad allergy phenotype (asthma, hay fever or eczema), including data from UK Biobank and 12 other contributing cohorts. | 180,129 cases with allergy and 180,709 controls. | <https://genepi.qimr.edu.au/staff/manuelF/gwas_results/main.html> |
| Smoking | Meta-analysis of ever smoker vs. never smoker with more than 30 contributing cohorts | 1.2 million individuals | <https://genome.psych.umn.edu/index.php/GSCAN> |

| **S4 Table.** SNPs used as instrumental variable for type 2 diabetes mellitus. | | | | | | | |
| --- | --- | --- | --- | --- | --- | --- | --- |
| **RS** | **CHR** | **EA** | **NEA** | **EAF** | **BETA** | **SE** | **PVAL** |
| rs1127655 | 1 | C | T | 0.470936254 | 0.0438 | 0.0079 | 2.47E-08 |
| rs12088739 | 1 | G | A | 0.089848136 | -0.0884 | 0.013 | 9.79E-12 |
| rs2296173 | 1 | G | A | 0.212010576 | 0.065 | 0.0087 | 7.66E-14 |
| rs2493394 | 1 | G | A | 0.107337868 | 0.073 | 0.0113 | 1.15E-10 |
| rs2820426 | 1 | A | G | 0.389942484 | -0.0521 | 0.0073 | 1.30E-12 |
| rs340874 | 1 | T | C | 0.436095755 | -0.0626 | 0.0073 | 8.41E-18 |
| rs348330 | 1 | G | A | 0.366548876 | 0.0487 | 0.0081 | 1.86E-09 |
| rs12617659 | 2 | T | C | 0.147238129 | -0.0685 | 0.0103 | 2.83E-11 |
| rs13389219 | 2 | T | C | 0.394367938 | -0.0722 | 0.0074 | 2.11E-22 |
| rs17334919 | 2 | T | C | 0.100217909 | -0.1398 | 0.0128 | 6.69E-28 |
| rs243019 | 2 | C | T | 0.455830717 | 0.0566 | 0.0071 | 2.29E-15 |
| rs2867125 | 2 | T | C | 0.172175282 | -0.0601 | 0.0096 | 4.33E-10 |
| rs2972144 | 2 | A | G | 0.354633108 | -0.0913 | 0.0075 | 2.55E-34 |
| rs7561798 | 2 | G | A | 0.482171025 | 0.04 | 0.0072 | 2.79E-08 |
| rs7572970 | 2 | A | G | 0.277953372 | -0.059 | 0.0087 | 1.39E-11 |
| rs780094 | 2 | T | C | 0.387154701 | -0.0692 | 0.0074 | 5.16E-21 |
| rs840967 | 2 | C | A | 0.394078203 | 0.0497 | 0.008 | 5.44E-10 |
| rs11708067 | 3 | G | A | 0.238989658 | -0.0965 | 0.0086 | 5.93E-29 |
| rs11925227 | 3 | A | G | 0.183438659 | -0.0534 | 0.0095 | 2.25E-08 |
| rs11926707 | 3 | T | C | 0.374443928 | -0.0463 | 0.0082 | 1.69E-08 |
| rs1496653 | 3 | G | A | 0.204781843 | -0.0769 | 0.0088 | 2.57E-18 |
| rs1899951 | 3 | T | C | 0.123288182 | -0.1118 | 0.0109 | 1.64E-24 |
| rs2292662 | 3 | T | C | 0.151271964 | -0.0629 | 0.0111 | 1.24E-08 |
| rs4686471 | 3 | T | C | 0.390224945 | -0.0534 | 0.0081 | 4.28E-11 |
| rs6795735 | 3 | T | C | 0.410911493 | -0.0558 | 0.0073 | 1.63E-14 |
| rs7651090 | 3 | G | A | 0.313376825 | 0.1204 | 0.0076 | 3.85E-57 |
| rs9844972 | 3 | C | G | 0.069722918 | 0.0956 | 0.0148 | 1.03E-10 |
| rs11098676 | 4 | T | C | 0.212360865 | -0.054 | 0.0096 | 2.03E-08 |
| rs17086692 | 4 | T | G | 0.313425861 | -0.0467 | 0.0084 | 2.48E-08 |
| rs1801214 | 4 | C | T | 0.400415373 | -0.0903 | 0.0074 | 5.52E-34 |
| rs735949 | 4 | C | T | 0.141149742 | -0.0711 | 0.0106 | 1.95E-11 |
| rs7674212 | 4 | T | G | 0.408864362 | -0.0465 | 0.0075 | 6.18E-10 |
| rs7685296 | 4 | T | C | 0.279364612 | -0.0511 | 0.0081 | 2.32E-10 |
| rs993380 | 4 | A | G | 0.334449877 | 0.0507 | 0.0081 | 4.59E-10 |
| rs10077431 | 5 | A | C | 0.21466813 | -0.0487 | 0.0089 | 4.76E-08 |
| rs1061813 | 5 | G | A | 0.462880802 | 0.0429 | 0.0073 | 3.37E-09 |
| rs459193 | 5 | A | G | 0.254662377 | -0.0711 | 0.0083 | 8.81E-18 |
| rs4865796 | 5 | G | A | 0.306909896 | -0.053 | 0.0078 | 1.33E-11 |
| rs6878122 | 5 | G | A | 0.318208635 | 0.0564 | 0.0079 | 1.19E-12 |
| rs7729395 | 5 | T | C | 0.050940934 | 0.1373 | 0.016 | 1.10E-17 |
| rs1050226 | 6 | G | A | 0.406792301 | -0.0491 | 0.0074 | 3.34E-11 |
| rs1063355 | 6 | T | G | 0.397563658 | -0.0709 | 0.0079 | 3.72E-19 |
| rs2246618 | 6 | T | C | 0.307253342 | 0.0513 | 0.0084 | 1.20E-09 |
| rs3756784 | 6 | G | T | 0.185838123 | 0.0505 | 0.0091 | 2.59E-08 |
| rs622217 | 6 | C | T | 0.483931554 | -0.0485 | 0.0077 | 3.13E-10 |
| rs72892910 | 6 | T | G | 0.172394288 | 0.0648 | 0.0099 | 6.43E-11 |
| rs7756992 | 6 | G | A | 0.266895681 | 0.1297 | 0.0078 | 6.00E-62 |
| rs853974 | 6 | T | C | 0.262413847 | 0.0601 | 0.0088 | 7.86E-12 |
| rs9369425 | 6 | G | A | 0.291815284 | 0.0546 | 0.0085 | 1.13E-10 |
| rs13239186 | 7 | T | C | 0.302028596 | 0.0539 | 0.0085 | 2.70E-10 |
| rs17168486 | 7 | T | C | 0.173603116 | 0.0742 | 0.0094 | 2.18E-15 |
| rs2191348 | 7 | G | T | 0.453064599 | -0.0652 | 0.0073 | 3.44E-19 |
| rs2299383 | 7 | T | C | 0.423455222 | 0.0412 | 0.0073 | 1.49E-08 |
| rs2908282 | 7 | A | G | 0.177394803 | 0.0552 | 0.0094 | 4.25E-09 |
| rs7786095 | 7 | G | A | 0.103859861 | -0.0743 | 0.0129 | 9.64E-09 |
| rs849135 | 7 | G | A | 0.50094782 | 0.0999 | 0.0072 | 1.04E-43 |
| rs10087241 | 8 | G | A | 0.405106648 | 0.0475 | 0.008 | 2.80E-09 |
| rs10100265 | 8 | A | C | 0.389510132 | 0.0491 | 0.0079 | 6.29E-10 |
| rs17411031 | 8 | G | C | 0.261729326 | -0.045 | 0.0081 | 3.04E-08 |
| rs2294120 | 8 | G | A | 0.4558793 | -0.0443 | 0.0079 | 1.62E-08 |
| rs3802177 | 8 | A | G | 0.311281439 | -0.1217 | 0.008 | 2.32E-52 |
| rs516946 | 8 | T | C | 0.239366701 | -0.0824 | 0.0085 | 3.16E-22 |
| rs7845219 | 8 | C | T | 0.492785851 | -0.0422 | 0.0072 | 4.55E-09 |
| rs10114341 | 9 | C | T | 0.440754141 | -0.0409 | 0.0072 | 1.15E-08 |
| rs10811661 | 9 | C | T | 0.173605396 | -0.1569 | 0.0098 | 4.13E-58 |
| rs10974438 | 9 | C | A | 0.351215241 | 0.0591 | 0.0075 | 3.01E-15 |
| rs1758632 | 9 | C | G | 0.376593207 | -0.0491 | 0.0081 | 1.36E-09 |
| rs17791483 | 9 | G | A | 0.062588323 | -0.102 | 0.0147 | 3.42E-12 |
| rs2796441 | 9 | A | G | 0.41645805 | -0.0715 | 0.0073 | 1.96E-22 |
| rs687621 | 9 | G | A | 0.324802052 | 0.0433 | 0.0076 | 1.35E-08 |
| rs10740322 | 10 | G | A | 0.31289619 | -0.0477 | 0.0085 | 2.11E-08 |
| rs11257655 | 10 | T | C | 0.206773281 | 0.0737 | 0.0087 | 1.97E-17 |
| rs35011184 | 10 | A | G | 0.227132731 | 0.2796 | 0.0088 | 1.00E-200 |
| rs753270 | 10 | T | C | 0.416465397 | -0.0528 | 0.0079 | 2.70E-11 |
| rs7923866 | 10 | T | C | 0.378774901 | -0.0972 | 0.0074 | 9.34E-40 |
| rs10830963 | 11 | G | C | 0.275768064 | 0.0909 | 0.008 | 5.85E-30 |
| rs1552224 | 11 | C | A | 0.154210453 | -0.1034 | 0.0101 | 8.64E-25 |
| rs2237892 | 11 | T | C | 0.062489595 | -0.096 | 0.0157 | 8.75E-10 |
| rs5215 | 11 | C | T | 0.360058778 | 0.0678 | 0.0073 | 2.09E-20 |
| rs67232546 | 11 | T | C | 0.209221793 | 0.0596 | 0.0096 | 4.66E-10 |
| rs7929543 | 11 | C | A | 0.083159887 | 0.0828 | 0.0138 | 2.20E-09 |
| rs10842994 | 12 | T | C | 0.197025395 | -0.0755 | 0.0091 | 1.02E-16 |
| rs11107116 | 12 | T | G | 0.219714311 | 0.0467 | 0.0085 | 3.75E-08 |
| rs12299509 | 12 | G | A | 0.478621745 | 0.0467 | 0.0073 | 2.09E-10 |
| rs2261181 | 12 | T | C | 0.096470033 | 0.0985 | 0.0118 | 9.18E-17 |
| rs61953351 | 12 | T | G | 0.249902348 | -0.07 | 0.0091 | 1.98E-14 |
| rs7138300 | 12 | C | T | 0.443165036 | 0.0443 | 0.0072 | 5.65E-10 |
| rs825476 | 12 | C | T | 0.419450711 | -0.0524 | 0.0073 | 6.81E-13 |
| rs1359790 | 13 | A | G | 0.286699864 | -0.0796 | 0.008 | 2.80E-23 |
| rs576674 | 13 | G | A | 0.167485416 | 0.0654 | 0.0097 | 1.79E-11 |
| rs963740 | 13 | T | A | 0.294299268 | -0.0479 | 0.0086 | 2.23E-08 |
| rs7144011 | 14 | T | G | 0.221063371 | 0.0482 | 0.0085 | 1.64E-08 |
| rs12910825 | 15 | G | A | 0.360390809 | 0.0517 | 0.0074 | 2.16E-12 |
| rs7177055 | 15 | G | A | 0.281711332 | -0.0647 | 0.0079 | 2.75E-16 |
| rs982077 | 15 | A | G | 0.434479032 | 0.0453 | 0.0072 | 2.58E-10 |
| rs13330951 | 16 | G | A | 0.488313929 | -0.0456 | 0.0081 | 1.54E-08 |
| rs2925979 | 16 | T | C | 0.299149856 | 0.0534 | 0.0078 | 9.06E-12 |
| rs7185735 | 16 | G | A | 0.397306062 | 0.1056 | 0.0073 | 1.59E-47 |
| rs77258096 | 16 | A | C | 0.101383462 | -0.1171 | 0.0134 | 1.78E-18 |
| rs9940149 | 16 | A | G | 0.178555946 | -0.058 | 0.0095 | 9.29E-10 |
| rs11651755 | 17 | C | T | 0.484669254 | 0.0741 | 0.0077 | 8.98E-22 |
| rs12945601 | 17 | T | C | 0.386396556 | 0.048 | 0.008 | 1.72E-09 |
| rs17405722 | 17 | A | G | 0.074152649 | 0.087 | 0.0146 | 2.28E-09 |
| rs17631783 | 17 | T | C | 0.263459811 | -0.0487 | 0.0089 | 3.95E-08 |
| rs8068804 | 17 | A | G | 0.325096537 | 0.0587 | 0.0078 | 4.41E-14 |
| rs9894220 | 17 | G | A | 0.433739993 | -0.0585 | 0.0079 | 1.52E-13 |
| rs12970134 | 18 | A | G | 0.265120386 | 0.0555 | 0.008 | 5.31E-12 |
| rs7240767 | 18 | C | T | 0.383677477 | 0.0451 | 0.0081 | 2.16E-08 |
| rs10401969 | 19 | C | T | 0.076585755 | 0.0921 | 0.0133 | 4.13E-12 |
| rs8108269 | 19 | G | T | 0.281015346 | 0.0644 | 0.0079 | 3.11E-16 |
| rs4810426 | 20 | T | C | 0.0967888 | 0.0726 | 0.013 | 2.15E-08 |
| rs6059662 | 20 | A | G | 0.336824454 | -0.0446 | 0.0079 | 1.51E-08 |
| rs6066138 | 20 | A | G | 0.278361954 | -0.049 | 0.0082 | 1.93E-09 |
| rs6515236 | 20 | C | A | 0.249330468 | -0.0504 | 0.0091 | 3.34E-08 |
| rs16988333 | 22 | G | A | 0.090403549 | -0.0745 | 0.013 | 9.17E-09 |
| rs4823182 | 22 | G | A | 0.335747643 | 0.0482 | 0.0077 | 3.36E-10 |
| CHR, chromosome; EA, effect allele; EAF, effect allele frequency; NEA, non-effect allele; SE, standard error; The beta coefficients represent one-unit increase in the log odds ratio of having type 2 diabetes for each additional effect allele. | | | | | | | |

| **S5 Table.** SNPs used as instrumental variable for coronary artery disease. | | | | | | | |
| --- | --- | --- | --- | --- | --- | --- | --- |
| **RS** | **CHR** | **EA** | **NEA** | **EAF** | **BETA** | **SE** | **PVAL** |
| rs10080815 | 6 | T | G | 0.972442 | -0.246627 | 0.0308579 | 1.33E-15 |
| rs10840293 | 11 | A | G | 0.549821 | 0.054714 | 0.009619 | 1.28E-08 |
| rs11065979 | 12 | C | T | 0.634501 | -0.068556 | 0.0107672 | 1.93E-10 |
| rs11191416 | 10 | T | G | 0.87253 | 0.079249 | 0.0135252 | 4.65E-09 |
| rs11556924 | 7 | C | T | 0.686675 | 0.072569 | 0.0110605 | 5.34E-11 |
| rs115654617 | 2 | C | A | 0.893038 | -0.137846 | 0.0158314 | 3.12E-18 |
| rs11838776 | 13 | G | A | 0.736723 | -0.068566 | 0.0107552 | 1.83E-10 |
| rs1199338 | 3 | A | C | 0.838134 | -0.073596 | 0.0124987 | 3.90E-09 |
| rs12202017 | 6 | A | G | 0.699953 | 0.066813 | 0.0099612 | 1.98E-11 |
| rs1412444 | 10 | C | T | 0.630869 | -0.066812 | 0.0096809 | 5.15E-12 |
| rs16986953 | 2 | G | A | 0.895294 | -0.08516 | 0.0150265 | 1.45E-08 |
| rs17087335 | 4 | G | T | 0.785363 | -0.060764 | 0.0111159 | 4.59E-08 |
| rs17678683 | 2 | T | G | 0.912319 | -0.098786 | 0.0166548 | 3.00E-09 |
| rs180803 | 22 | T | G | 0.029268 | -0.180923 | 0.0283062 | 1.64E-10 |
| rs1870634 | 10 | G | T | 0.637485 | 0.075878 | 0.0097113 | 5.55E-15 |
| rs2107595 | 7 | G | A | 0.79953 | -0.073415 | 0.0112951 | 8.05E-11 |
| rs2128739 | 11 | C | A | 0.676464 | -0.065565 | 0.0100568 | 7.05E-11 |
| rs2487928 | 10 | G | A | 0.581779 | -0.062633 | 0.0095049 | 4.41E-11 |
| rs2519093 | 9 | C | T | 0.809128 | -0.079704 | 0.0117524 | 1.19E-11 |
| rs2681472 | 12 | A | G | 0.798694 | -0.074114 | 0.0113331 | 6.17E-11 |
| rs28451064 | 21 | G | A | 0.878814 | -0.127571 | 0.015952 | 1.33E-15 |
| rs2891168 | 9 | A | G | 0.511332 | -0.193401 | 0.0091877 | 2.29E-98 |
| rs3918226 | 7 | C | T | 0.935485 | -0.133315 | 0.0221275 | 1.69E-09 |
| rs4420638 | 19 | A | G | 0.833964 | -0.091906 | 0.0140977 | 7.07E-11 |
| rs4468572 | 15 | C | T | 0.585831 | 0.077234 | 0.0095277 | 4.44E-16 |
| rs4593108 | 4 | C | G | 0.795349 | 0.07083 | 0.0115558 | 8.82E-10 |
| rs515135 | 2 | C | T | 0.791985 | 0.067499 | 0.0121924 | 3.09E-08 |
| rs55730499 | 6 | C | T | 0.943757 | -0.316641 | 0.0242403 | 5.39E-39 |
| rs56062135 | 15 | C | T | 0.794271 | 0.069743 | 0.0118937 | 4.52E-09 |
| rs56289821 | 19 | G | A | 0.899622 | 0.13361 | 0.0170415 | 4.44E-15 |
| rs56336142 | 6 | T | C | 0.807262 | 0.066813 | 0.0118763 | 1.85E-08 |
| rs663129 | 18 | G | A | 0.743165 | -0.058163 | 0.0105173 | 3.20E-08 |
| rs6689306 | 1 | G | A | 0.552455 | -0.056012 | 0.0094061 | 2.60E-09 |
| rs67180937 | 1 | G | T | 0.663052 | 0.078807 | 0.0110551 | 1.01E-12 |
| rs7212798 | 17 | T | C | 0.853484 | -0.079961 | 0.0142216 | 1.88E-08 |
| rs7528419 | 1 | A | G | 0.78582 | 0.11453 | 0.011482 | 1.97E-23 |
| rs8042271 | 15 | A | G | 0.097718 | -0.096711 | 0.0175662 | 3.68E-08 |
| rs9349379 | 6 | A | G | 0.568394 | -0.131836 | 0.0096527 | 1.81E-42 |
| rs9970807 | 1 | C | T | 0.915097 | 0.12575 | 0.016695 | 5.00E-14 |
| CHR, chromosome; EA, effect allele; EAF, effect allele frequency; NEA, non-effect allele; SE, standard error; The beta coefficients represent one-unit increase in the log odds ratio of having coronary artery disease for each additional effect allele. | | | | | | | |

| **S6 Table.** SNPs used as instrumental variable for allergy. | | | | | | | |
| --- | --- | --- | --- | --- | --- | --- | --- |
| **RS** | **CHR** | **EA** | **NEA** | **EAF** | **BETA** | **SE** | **PVAL** |
| rs10033073 | 4 | A | G | 0.3503 | -0.0437 | 0.0072 | 1.36E-09 |
| rs10174949 | 2 | A | G | 0.3163 | -0.0656 | 0.0063 | 9.92E-26 |
| rs10414065 | 19 | T | C | 0.07823 | -0.0917 | 0.012 | 2.29E-14 |
| rs10519067 | 15 | A | G | 0.1395 | -0.0518 | 0.0085 | 1.10E-09 |
| rs1059513 | 12 | T | C | 0.1105 | 0.0828 | 0.0094 | 1.15E-18 |
| rs10789841 | 11 | T | C | 0.3095 | 0.0456 | 0.0064 | 1.48E-12 |
| rs10865050 | 2 | A | G | 0.1531 | -0.1249 | 0.0084 | 6.37E-50 |
| rs11033545 | 11 | T | G | 0.4422 | -0.0334 | 0.006 | 2.41E-08 |
| rs11236814 | 11 | A | T | 0.09184 | 0.0653 | 0.0098 | 3.25E-11 |
| rs12123821 | 1 | T | C | 0.04762 | 0.1159 | 0.014 | 1.04E-16 |
| rs1214598 | 1 | A | G | 0.3316 | -0.0401 | 0.006 | 2.14E-11 |
| rs12365699 | 11 | A | G | 0.1531 | -0.0592 | 0.0078 | 3.54E-14 |
| rs12413578 | 10 | T | C | 0.09864 | -0.0934 | 0.0094 | 3.31E-23 |
| rs12440045 | 15 | A | C | 0.4524 | -0.0403 | 0.0059 | 7.68E-12 |
| rs12551834 | 9 | A | G | 0.07993 | -0.0615 | 0.0103 | 2.72E-09 |
| rs12625547 | 20 | T | G | 0.2143 | 0.0432 | 0.0077 | 1.98E-08 |
| rs1289273 | 1 | A | G | 0.4269 | -0.035 | 0.0057 | 1.09E-09 |
| rs12941864 | 17 | T | C | 0.466 | -0.0335 | 0.0061 | 4.98E-08 |
| rs1419675 | 6 | T | G | 0.2415 | -0.0385 | 0.007 | 4.72E-08 |
| rs144829310 | 9 | T | G | 0.1565 | 0.0828 | 0.0078 | 2.60E-26 |
| rs1555926 | 20 | T | C | 0.2041 | -0.0543 | 0.0072 | 5.58E-14 |
| rs1689510 | 12 | C | G | 0.3163 | 0.051 | 0.0061 | 3.39E-17 |
| rs16903574 | 5 | C | G | 0.08163 | -0.07 | 0.011 | 1.68E-10 |
| rs17664743 | 7 | A | G | 0.2041 | 0.041 | 0.0071 | 8.27E-09 |
| rs1885013 | 14 | A | G | 0.3588 | -0.0382 | 0.0063 | 1.33E-09 |
| rs2134814 | 6 | C | G | 0.3537 | 0.0427 | 0.006 | 1.03E-12 |
| rs2212434 | 11 | T | C | 0.4286 | 0.0869 | 0.0057 | 8.93E-52 |
| rs2221641 | 8 | T | C | 0.3878 | -0.0419 | 0.0059 | 1.03E-12 |
| rs2241099 | 16 | C | G | 0.2619 | 0.0697 | 0.0066 | 4.62E-26 |
| rs228619 | 4 | A | G | 0.4694 | -0.0362 | 0.0057 | 2.41E-10 |
| rs2477923 | 10 | T | C | 0.4983 | 0.0332 | 0.0057 | 7.23E-09 |
| rs249677 | 5 | A | C | 0.3605 | 0.0366 | 0.0059 | 7.54E-10 |
| rs2854001 | 6 | A | G | 0.1837 | 0.0537 | 0.0071 | 3.31E-14 |
| rs2910162 | 5 | A | G | 0.3163 | -0.0344 | 0.0061 | 1.54E-08 |
| rs3024665 | 16 | T | C | 0.07653 | 0.0665 | 0.0119 | 2.18E-08 |
| rs3128959 | 6 | A | G | 0.1241 | -0.0601 | 0.0095 | 2.25E-10 |
| rs34004019 | 6 | A | G | 0.3044 | 0.0913 | 0.0071 | 2.52E-38 |
| rs3540 | 15 | A | G | 0.3452 | -0.0346 | 0.0061 | 1.28E-08 |
| rs4296977 | 7 | T | C | 0.1582 | -0.0534 | 0.0082 | 6.92E-11 |
| rs479844 | 11 | A | G | 0.4286 | -0.0412 | 0.0058 | 1.15E-12 |
| rs4943794 | 13 | C | G | 0.2262 | 0.0396 | 0.007 | 1.41E-08 |
| rs4973380 | 2 | A | T | 0.2432 | 0.0397 | 0.0067 | 2.59E-09 |
| rs519973 | 3 | A | G | 0.3537 | 0.0358 | 0.006 | 3.23E-09 |
| rs56375023 | 15 | A | G | 0.2126 | 0.0731 | 0.0068 | 3.11E-27 |
| rs56389811 | 12 | T | C | 0.199 | -0.0464 | 0.007 | 3.13E-11 |
| rs5743618 | 4 | A | C | 0.2993 | -0.0915 | 0.0067 | 2.73E-42 |
| rs5758343 | 22 | A | T | 0.2177 | 0.0474 | 0.0071 | 2.17E-11 |
| rs58939053 | 4 | T | C | 0.3435 | 0.0544 | 0.0061 | 3.85E-19 |
| rs6011033 | 20 | A | G | 0.2245 | -0.0437 | 0.0069 | 2.66E-10 |
| rs61192126 | 3 | T | C | 0.3095 | 0.0384 | 0.0064 | 1.56E-09 |
| rs61816766 | 1 | T | C | 0.02041 | -0.142 | 0.0176 | 6.76E-16 |
| rs62626322 | 10 | T | G | 0.09694 | -0.0674 | 0.0091 | 1.35E-13 |
| rs6461503 | 7 | T | C | 0.4728 | 0.041 | 0.0057 | 8.81E-13 |
| rs6489785 | 12 | T | C | 0.369 | 0.0428 | 0.0059 | 3.57E-13 |
| rs6594499 | 5 | A | C | 0.4813 | -0.0723 | 0.0057 | 1.10E-36 |
| rs6800001 | 3 | A | G | 0.3418 | 0.033 | 0.0059 | 2.28E-08 |
| rs6881706 | 5 | T | G | 0.2925 | -0.0707 | 0.0064 | 1.73E-28 |
| rs6990534 | 8 | A | G | 0.3571 | 0.04 | 0.0063 | 1.74E-10 |
| rs7224129 | 17 | A | G | 0.4728 | 0.0535 | 0.0057 | 8.11E-21 |
| rs72774901 | 5 | A | T | 0.08163 | 0.1122 | 0.0108 | 3.92E-25 |
| rs7406234 | 17 | T | C | 0.4354 | 0.0337 | 0.0057 | 4.30E-09 |
| rs74847330 | 2 | A | G | 0.1224 | 0.0497 | 0.0088 | 1.67E-08 |
| rs7521390 | 1 | A | C | 0.284 | -0.0358 | 0.0063 | 1.40E-08 |
| rs7625643 | 3 | A | G | 0.4422 | -0.0346 | 0.0058 | 2.96E-09 |
| rs7712601 | 5 | T | C | 0.4184 | 0.0346 | 0.0058 | 3.28E-09 |
| rs80064395 | 3 | T | C | 0.05782 | -0.0726 | 0.0107 | 1.32E-11 |
| rs8030821 | 15 | A | T | 0.3895 | -0.0337 | 0.006 | 1.54E-08 |
| rs848 | 5 | A | C | 0.2364 | 0.0601 | 0.0073 | 1.62E-16 |
| rs9372120 | 6 | T | G | 0.1752 | -0.0406 | 0.0071 | 1.02E-08 |
| rs9877752 | 3 | A | G | 0.4116 | 0.0398 | 0.0058 | 4.98E-12 |
| CHR, chromosome; EA, effect allele; EAF, effect allele frequency; NEA, non-effect allele; SE, standard error; The beta coefficients represent one-unit increase in the log odds ratio of having allergy for each additional effect allele. | | | | | | | |

| **S7 Table.** SNPs used as instrumental variable for smoking. | | | | | | | |
| --- | --- | --- | --- | --- | --- | --- | --- |
| **RS** | **CHR** | **EA** | **NEA** | **EAF** | **BETA** | **SE** | **PVAL** |
| rs10001365 | 4 | G | A | 0.405 | -0.0249918 | 0.00364155 | 6.65E-12 |
| rs1004787 | 2 | G | A | 0.581 | 0.02992312 | 0.0035714 | 5.27E-17 |
| rs10114490 | 9 | G | A | 0.198 | -0.0255148 | 0.00453171 | 1.81E-08 |
| rs10233018 | 7 | A | G | 0.503 | 0.02706904 | 0.00355741 | 2.75E-14 |
| rs10260968 | 7 | G | A | 0.597 | -0.0203218 | 0.00360938 | 1.75E-08 |
| rs10279261 | 7 | G | A | 0.619 | -0.0214194 | 0.00366263 | 5.00E-09 |
| rs10498846 | 6 | C | T | 0.473 | 0.02061029 | 0.00355561 | 6.62E-09 |
| rs1050847 | 16 | C | T | 0.505 | -0.0216231 | 0.00358893 | 1.67E-09 |
| rs10828246 | 10 | T | C | 0.385 | 0.02492553 | 0.00369927 | 1.61E-11 |
| rs10905461 | 10 | T | C | 0.718 | -0.0239554 | 0.00414505 | 7.35E-09 |
| rs11057005 | 12 | A | G | 0.43 | -0.0209298 | 0.00357892 | 4.85E-09 |
| rs11078713 | 17 | A | G | 0.454 | -0.0201721 | 0.00360561 | 2.23E-08 |
| rs1154693 | 3 | A | G | 0.856 | 0.03262167 | 0.00491232 | 3.12E-11 |
| rs11658881 | 17 | A | G | 0.418 | 0.02013572 | 0.00361066 | 2.43E-08 |
| rs11712680 | 3 | A | C | 0.174 | -0.0270476 | 0.00457843 | 3.51E-09 |
| rs117143374 | 21 | T | C | 0.12 | 0.02928969 | 0.00526909 | 2.76E-08 |
| rs11721059 | 3 | C | T | 0.474 | 0.01993569 | 0.00356335 | 2.17E-08 |
| rs12027999 | 1 | T | C | 0.124 | -0.0330855 | 0.00533915 | 5.76E-10 |
| rs12042107 | 1 | T | C | 0.527 | -0.0222834 | 0.0035682 | 4.22E-10 |
| rs12112638 | 7 | A | G | 0.275 | -0.024526 | 0.00404299 | 1.34E-09 |
| rs12186738 | 5 | G | T | 0.154 | -0.0332644 | 0.00502051 | 3.42E-11 |
| rs12333760 | 7 | T | C | 0.204 | -0.0290467 | 0.00480127 | 1.44E-09 |
| rs12356821 | 10 | G | C | 0.14 | 0.03937004 | 0.0050491 | 6.27E-15 |
| rs12441907 | 15 | C | A | 0.186 | -0.0292051 | 0.00452262 | 1.06E-10 |
| rs12474587 | 2 | G | T | 0.404 | 0.02763286 | 0.00358234 | 1.25E-14 |
| rs12545053 | 8 | A | G | 0.397 | 0.0202808 | 0.00363668 | 2.43E-08 |
| rs12632110 | 3 | A | G | 0.647 | -0.0233768 | 0.00375292 | 4.78E-10 |
| rs12923427 | 16 | C | T | 0.204 | -0.0239087 | 0.0043724 | 4.44E-08 |
| rs13030994 | 2 | G | A | 0.485 | 0.03609247 | 0.0035563 | 3.56E-24 |
| rs13145728 | 4 | G | C | 0.358 | -0.0232512 | 0.00366263 | 2.14E-10 |
| rs13261666 | 8 | G | T | 0.522 | -0.0268946 | 0.00355604 | 3.90E-14 |
| rs134529 | 22 | T | C | 0.349 | -0.019984 | 0.00366078 | 4.85E-08 |
| rs1385108 | 5 | C | T | 0.239 | 0.0246617 | 0.00415673 | 3.00E-09 |
| rs1435741 | 15 | G | A | 0.425 | 0.02941512 | 0.00359095 | 2.64E-16 |
| rs1445649 | 2 | T | C | 0.525 | 0.02399323 | 0.00356484 | 1.68E-11 |
| rs1518393 | 2 | A | C | 0.631 | 0.02053574 | 0.00365894 | 2.03E-08 |
| rs1555445 | 20 | A | T | 0.337 | 0.0225548 | 0.0038234 | 3.65E-09 |
| rs1565735 | 8 | T | A | 0.212 | -0.037618 | 0.0044613 | 3.42E-17 |
| rs1899896 | 8 | C | T | 0.286 | 0.02644812 | 0.00388691 | 1.04E-11 |
| rs1971318 | 12 | C | T | 0.141 | 0.02850742 | 0.00492533 | 7.06E-09 |
| rs2046850 | 1 | C | T | 0.187 | -0.0248139 | 0.00447842 | 3.03E-08 |
| rs2050586 | 1 | G | C | 0.355 | -0.0205467 | 0.00370828 | 3.00E-08 |
| rs2107300 | 2 | C | G | 0.845 | -0.027201 | 0.00492533 | 3.27E-08 |
| rs222449 | 6 | A | T | 0.793 | -0.0253208 | 0.00442796 | 1.08E-08 |
| rs2378662 | 9 | G | A | 0.556 | 0.02094815 | 0.00356645 | 4.16E-09 |
| rs240963 | 6 | T | C | 0.836 | -0.0410444 | 0.00483712 | 2.16E-17 |
| rs266047 | 2 | G | A | 0.529 | -0.0305098 | 0.00373855 | 3.36E-16 |
| rs292071 | 4 | T | C | 0.244 | 0.0235822 | 0.00400909 | 4.08E-09 |
| rs3001723 | 1 | G | A | 0.321 | 0.0335118 | 0.0038983 | 8.12E-18 |
| rs301805 | 1 | T | G | 0.559 | 0.02146792 | 0.00361329 | 2.80E-09 |
| rs35702515 | 2 | G | T | 0.162 | 0.02524417 | 0.00423093 | 2.43E-09 |
| rs3800227 | 6 | A | G | 0.701 | 0.02281212 | 0.00405809 | 1.93E-08 |
| rs3801289 | 7 | A | C | 0.351 | -0.0220618 | 0.00373982 | 3.74E-09 |
| rs3904512 | 13 | G | A | 0.429 | -0.0211589 | 0.0035765 | 3.23E-09 |
| rs4044321 | 5 | A | G | 0.642 | -0.0278417 | 0.00371058 | 6.08E-14 |
| rs4236259 | 7 | T | G | 0.499 | -0.0247689 | 0.00355661 | 3.35E-12 |
| rs4523689 | 11 | A | G | 0.408 | -0.0206091 | 0.00364321 | 1.55E-08 |
| rs4543592 | 9 | T | C | 0.468 | 0.02193143 | 0.00356244 | 7.46E-10 |
| rs4571506 | 5 | C | T | 0.492 | -0.0275747 | 0.00356881 | 1.09E-14 |
| rs4674993 | 2 | A | G | 0.207 | -0.0252122 | 0.00443619 | 1.32E-08 |
| rs4759228 | 12 | G | C | 0.27 | -0.0216913 | 0.00393413 | 3.58E-08 |
| rs4785836 | 16 | T | C | 0.398 | -0.0204704 | 0.00365894 | 2.26E-08 |
| rs56820925 | 20 | C | T | 0.347 | -0.0218638 | 0.00387715 | 1.73E-08 |
| rs6265 | 11 | C | T | 0.203 | -0.0317863 | 0.00457843 | 3.77E-12 |
| rs6433897 | 2 | T | C | 0.754 | 0.02244827 | 0.00405809 | 3.16E-08 |
| rs6565846 | 18 | A | G | 0.722 | 0.02212049 | 0.00401859 | 3.77E-08 |
| rs6669839 | 1 | C | T | 0.204 | 0.02600412 | 0.0043955 | 3.36E-09 |
| rs6728726 | 2 | T | C | 0.829 | 0.03544859 | 0.00473279 | 6.73E-14 |
| rs6788098 | 3 | A | T | 0.623 | -0.0313461 | 0.00368905 | 1.91E-17 |
| rs6893752 | 5 | A | G | 0.766 | -0.0240995 | 0.00407356 | 3.25E-09 |
| rs7197072 | 16 | C | T | 0.238 | -0.0247672 | 0.0041686 | 2.77E-09 |
| rs7224742 | 17 | C | T | 0.595 | -0.0207099 | 0.00365532 | 1.43E-08 |
| rs72789632 | 5 | C | T | 0.12 | -0.0328856 | 0.00528628 | 5.02E-10 |
| rs72896886 | 18 | G | C | 0.144 | -0.0268885 | 0.00483712 | 2.75E-08 |
| rs7322872 | 13 | C | T | 0.782 | -0.0255713 | 0.00433474 | 3.58E-09 |
| rs7555507 | 1 | C | T | 0.496 | -0.0241444 | 0.00355604 | 1.14E-11 |
| rs76214862 | 14 | A | C | 0.202 | -0.0249903 | 0.00454745 | 3.99E-08 |
| rs7631735 | 3 | G | T | 0.603 | 0.0202273 | 0.00364471 | 2.79E-08 |
| rs76608582 | 19 | C | A | 0.0389 | -0.0495575 | 0.00825958 | 1.94E-09 |
| rs7929518 | 11 | A | G | 0.765 | 0.02423769 | 0.00428466 | 1.56E-08 |
| rs7938812 | 11 | T | G | 0.424 | 0.0437914 | 0.00363668 | 2.71E-33 |
| rs7969559 | 12 | A | G | 0.688 | -0.0243756 | 0.00395946 | 7.31E-10 |
| rs9401770 | 6 | G | A | 0.273 | 0.02773066 | 0.003986 | 3.47E-12 |
| rs993700 | 4 | T | C | 0.766 | -0.025928 | 0.00429163 | 1.53E-09 |
| CHR, chromosome; EA, effect allele; EAF, effect allele frequency; NEA, non-effect allele; SE, standard error; The beta coefficients represent one-unit increase in the log odds ratio of having coronary artery disease for each additional effect allele. | | | | | | | |

| **S8 Table.** Participant characteristics of ACE inhibitor-associated angioedema cases and ACE inhibitor-exposed controls in the SwedeGene cohort. | | |
| --- | --- | --- |
|  | **Cases** | **Controls** |
|  | *n=144* | *n=1345* |
| Age, mean (range) | 66.4 [31, 91] | 68.9 [49–96] |
| Male sex, *N* (%) | 250 (54.1) | 864 (64.0) |
| Medical conditions, *N* (%) |  |  |
| Hypertension | 134 (93.1) | 668 (69.9) |
| Diabetes | 35 (24.3) | 230 (24.1) |
| Heart failure | 5 (3.5) | 162 (16.9) |
| Type of ACEi, *N* (%) |  |  |
| C09AA01, Captopril | 0 (0) | 13 (1.0) |
| C09AA02, Enalapril | 116 (80.6) | 1055 (78.4) |
| C09AA03, Lisinopril | 3 (2.1) | 27 (2.0) |
| C09AA05, Ramipril | 24 (16.7) | 239 (17.8) |
| C09AA06, Quinapril | 1 (0.7) | 1 (0.1) |
| C09AA08 Cilazapril | 0 (0) | 8 (0.6) |
| C09AA09, Fosinopril | 0 (0.0) | 1 (0.1) |
| C09AA10, Trandolapril | 37 (8.0) | 1 (0.1) |

| **S9 Table.** Variants linked with ACEi-associated angioedema in the literature | | | | | | | | | |
| --- | --- | --- | --- | --- | --- | --- | --- | --- | --- |
| **Variant** | **Chr:Pos** | **Effect allele** | **Other allele** | **EAF** | **Gene** | **Beta** | **SE** | **Pval** | **PMID** |
| rs6020 | 1:169519112 | T | C | 0.01 | F5 | 0.080 | 0.541 | 0.883 | 32496628 |
| rs989692 | 3:154801365 | T | C | 0.49 | *MME* | -0.155 | 0.128 | 0.224 | 23838604 |
| rs2253201 | 10:79356397 | G | A | 0.05 | *KCNMA1* | -0.015 | 0.147 | 0.919 | 32080354 |
| rs500766 | 10:6550590 | T | C | 0.27 | *PRKCQ* | -0.045 | 0.066 | 0.493 | 23838604 |
| rs2724635 | [12:11899973](https://grch37.ensembl.org/Homo_sapiens/Location/View?contigviewbottom=variation_feature_variation%3Dnormal%2Cseq%3Dnormal;db=core;r=12:11899923-11900023;source=dbSNP;v=rs2724635;vdb=variation;vf=248796398) | G | A | 0.50 | *ETV6* | 0.065 | 0.060 | 0.280 | 23838604 |

| **S10 Table.** Summary statistics and functional annotation for SNPs reaching genome-wide significance in a meta-analysis of ACE inhibitor associated angioedema | | | | | | | | | | | | | |  |
| --- | --- | --- | --- | --- | --- | --- | --- | --- | --- | --- | --- | --- | --- | --- |
| **Sentinel variant** | **Chromosome** | **hg19 position** | **Non effect allele** | **Effect allele** | **Minor allele frequency** | **LD *r^2^ with lead SNP*** | ***P*-value** | **Beta** | **SE** | **Nearest Gene** | **Location** | **CADD** | **RDB** | **minChrState** |
| rs72704813 | 14 | 96612609 | G | A | 0.2286 | 1.00 | 1.65E-10 | -0.4744 | 0.0742 | BDKRB2 | intergenic | 18.11 | 2b | 2 |
| rs12885218 | 14 | 96614325 | G | A | 0.2485 | 0.89 | 1.27E-08 | -0.4057 | 0.0713 | BDKRB2 | intergenic | 9.589 | 7 | 5 |
| rs11850332 | 14 | 96618733 | G | C | 0.7664 | 0.97 | 2.06E-10 | 0.4708 | 0.0741 | BDKRB2 | intergenic | 8.771 | 6 | 5 |
| rs12894970 | 14 | 96612682 | C | T | 0.7505 | 0.89 | 1.05E-08 | 0.4079 | 0.0713 | BDKRB2 | intergenic | 7.988 | 4 | 2 |
| rs11850303 | 14 | 96618740 | C | A | 0.7664 | 0.97 | 2.06E-10 | 0.4708 | 0.0741 | BDKRB2 | intergenic | 7.709 | 7 | 5 |
| rs10140368 | 14 | 96616571 | G | A | 0.7515 | 0.89 | 1.20E-08 | 0.4063 | 0.0713 | BDKRB2 | intergenic | 7.181 | 6 | 5 |
| rs11846465 | 14 | 96620639 | G | A | 0.7664 | 0.97 | 1.65E-10 | 0.4735 | 0.0741 | BDKRB2 | intergenic | 5.994 | 7 | 5 |
| rs34845487 | 14 | 96617046 | C | A | 0.2286 | 1.00 | 2.01E-10 | -0.472 | 0.0742 | BDKRB2 | intergenic | 5.62 | 5 | 5 |
| rs7492727 | 14 | 96622909 | C | A | 0.2495 | 0.89 | 1.30E-08 | -0.4054 | 0.0713 | BDKRB2 | intergenic | 5.101 | 6 | 5 |
| rs11846550 | 14 | 96620826 | G | A | 0.7664 | 0.97 | 1.66E-10 | 0.4735 | 0.0741 | BDKRB2 | intergenic | 4.674 | 7 | 5 |
| rs71415028 | 14 | 96620955 | G | A | 0.7664 | 0.97 | 1.66E-10 | 0.4735 | 0.0741 | BDKRB2 | intergenic | 4.221 | 7 | 5 |
| rs68023675 | 14 | 96615137 | G | A | 0.7714 | 1.00 | 1.68E-10 | 0.4741 | 0.0742 | BDKRB2 | intergenic | 3.866 | 2c | 5 |
| rs11846417 | 14 | 96620556 | G | A | 0.7664 | 0.97 | 1.67E-10 | 0.4734 | 0.0741 | BDKRB2 | intergenic | 3.777 | 6 | 5 |
| rs11846531 | 14 | 96620508 | G | A | 0.2336 | 0.97 | 1.66E-10 | -0.4735 | 0.0741 | BDKRB2 | intergenic | 3.551 | 7 | 5 |
| rs71415027 | 14 | 96620874 | C | T | 0.2336 | 0.97 | 1.65E-10 | -0.4735 | 0.0741 | BDKRB2 | intergenic | 3.398 | 7 | 5 |
| rs11846378 | 14 | 96620441 | G | A | 0.7664 | 0.97 | 1.65E-10 | 0.4735 | 0.0741 | BDKRB2 | intergenic | 3.385 | 7 | 5 |
| rs35136400 | 14 | 96619480 | G | A | 0.7664 | 0.97 | 1.64E-10 | 0.4736 | 0.0741 | BDKRB2 | intergenic | 3.28 | 5 | 5 |
| rs66680728 | 14 | 96625772 | C | T | 0.7654 | 0.97 | 1.50E-10 | 0.4755 | 0.0742 | BDKRB2 | intergenic | 3.265 | 7 | 5 |
| rs34393530 | 14 | 96624517 | G | A | 0.7694 | 0.99 | 1.37E-10 | 0.4772 | 0.0743 | BDKRB2 | intergenic | 3.247 | 7 | 5 |
| rs2369544 | 14 | 96621379 | G | A | 0.7664 | 0.97 | 1.68E-10 | 0.4734 | 0.0741 | BDKRB2 | intergenic | 3.245 | 6 | 5 |
| rs2369541 | 14 | 96600784 | G | T | 0.7724 | 0.98 | 3.28E-10 | 0.4665 | 0.0742 | BDKRB2 | intergenic | 3.111 | 7 | 5 |
| rs12888576 | 14 | 96611391 | G | A | 0.7714 | 1.00 | 1.02E-10 | 0.4804 | 0.0743 | BDKRB2 | intergenic | 2.981 | 5 | 5 |
| rs59804216 | 14 | 96600316 | C | T | 0.2286 | 0.98 | 3.83E-10 | -0.4651 | 0.0743 | BDKRB2 | intergenic | 2.771 | 5 | 5 |
| rs71415026 | 14 | 96615089 | G | A | 0.7714 | 1.00 | 1.68E-10 | 0.4741 | 0.0742 | BDKRB2 | intergenic | 2.755 | 4 | 5 |
| rs4627266 | 14 | 96600596 | T | A | 0.2276 | 0.98 | 3.77E-10 | -0.4652 | 0.0743 | BDKRB2 | intergenic | 2.684 | 6 | 5 |
| rs34485356 | 14 | 96611271 | C | T | 0.7664 | 1.00 | 1.08E-10 | 0.4797 | 0.0743 | BDKRB2 | intergenic | 2.365 | 6 | 5 |
| rs1959041 | 14 | 96617880 | G | A | 0.7664 | 0.89 | 1.45E-08 | 0.4037 | 0.0712 | BDKRB2 | intergenic | 2.333 | *NA* | 5 |
| rs12894873 | 14 | 96619739 | G | T | 0.2336 | 0.97 | 1.64E-10 | -0.4736 | 0.0741 | BDKRB2 | intergenic | 2.307 | 7 | 5 |
| rs11850334 | 14 | 96618781 | T | A | 0.7664 | 0.97 | 2.05E-10 | 0.4708 | 0.0741 | BDKRB2 | intergenic | 2.15 | 7 | 5 |
| rs4905449 | 14 | 96616330 | G | T | 0.7505 | 0.89 | 1.17E-08 | 0.4066 | 0.0713 | BDKRB2 | intergenic | 1.852 | 7 | 5 |
| rs34033283 | 14 | 96599656 | G | A | 0.2276 | 0.97 | 1.09E-09 | -0.454 | 0.0745 | BDKRB2 | intergenic | 1.78 | 7 | 5 |
| rs11160314 | 14 | 96604435 | C | T | 0.7565 | 0.91 | 2.09E-08 | 0.399 | 0.0712 | BDKRB2 | intergenic | 1.667 | 7 | 5 |
| rs55940712 | 14 | 96605573 | G | A | 0.2286 | 0.99 | 2.17E-10 | -0.4716 | 0.0743 | BDKRB2 | intergenic | 1.655 | 5 | 7 |
| rs35974883 | 14 | 96617205 | G | A | 0.2336 | 0.97 | 2.88E-10 | -0.4667 | 0.074 | BDKRB2 | intergenic | 1.615 | 7 | 5 |
| rs1889372 | 14 | 96603815 | G | C | 0.2435 | 0.91 | 2.07E-08 | -0.3991 | 0.0712 | BDKRB2 | downstream | 1.545 | 5 | 5 |
| rs34985854 | 14 | 96617940 | C | T | 0.2286 | 1.00 | 1.62E-10 | -0.4747 | 0.0742 | BDKRB2 | intergenic | 1.534 | 7 | 5 |
| rs36092996 | 14 | 96616039 | G | C | 0.7664 | 0.97 | 1.64E-10 | 0.4737 | 0.0741 | BDKRB2 | intergenic | 1.468 | 7 | 5 |
| rs34870532 | 14 | 96624719 | G | T | 0.2296 | 0.99 | 1.50E-10 | -0.4763 | 0.0744 | BDKRB2 | intergenic | 1.398 | 7 | 5 |
| rs55668608 | 14 | 96619307 | G | T | 0.2336 | 0.97 | 1.64E-10 | -0.4736 | 0.0741 | BDKRB2 | intergenic | 1.396 | 7 | 5 |
| rs12881275 | 14 | 96622754 | G | A | 0.2505 | 0.89 | 1.30E-08 | -0.4055 | 0.0713 | BDKRB2 | intergenic | 1.329 | 6 | 5 |
| rs12432014 | 14 | 96622427 | C | T | 0.2485 | 0.89 | 1.33E-08 | -0.4052 | 0.0713 | BDKRB2 | intergenic | 1.183 | 6 | 5 |
| rs4905447 | 14 | 96602783 | G | A | 0.7565 | 0.91 | 2.04E-08 | 0.3992 | 0.0712 | BDKRB2 | upstream | 1.112 | 2b | 5 |
| rs28823359 | 14 | 96621833 | G | C | 0.7664 | 0.97 | 1.68E-10 | 0.4734 | 0.0741 | BDKRB2 | intergenic | 1.019 | 7 | 5 |
| rs28849215 | 14 | 96621809 | G | A | 0.2336 | 0.97 | 1.67E-10 | -0.4734 | 0.0741 | BDKRB2 | intergenic | 0.97 | 6 | 5 |
| rs2151767 | 14 | 96608962 | C | T | 0.2276 | 0.99 | 1.93E-10 | -0.4729 | 0.0743 | BDKRB2 | intergenic | 0.927 | *NA* | 5 |
| rs11850248 | 14 | 96618312 | G | A | 0.2336 | 0.97 | 2.11E-10 | -0.4705 | 0.0741 | BDKRB2 | intergenic | 0.793 | 7 | 5 |
| rs2369539 | 14 | 96600583 | C | T | 0.2276 | 0.98 | 5.48E-10 | -0.4604 | 0.0742 | BDKRB2 | intergenic | 0.707 | 6 | 5 |
| rs8022837 | 14 | 96616845 | C | T | 0.2336 | 0.97 | 2.62E-10 | -0.4677 | 0.074 | BDKRB2 | intergenic | 0.554 | 6 | 5 |
| rs7156430 | 14 | 96603267 | C | T | 0.7565 | 0.91 | 2.09E-08 | 0.399 | 0.0712 | BDKRB2 | ncRNA_exonic | 0.551 | 5 | 7 |
| rs60634508 | 14 | 96606733 | C | T | 0.2296 | 0.98 | 1.55E-10 | -0.4758 | 0.0743 | BDKRB2 | intergenic | 0.483 | 5 | 5 |
| rs3939400 | 14 | 96604627 | C | T | 0.2435 | 0.91 | 2.29E-08 | -0.398 | 0.0712 | BDKRB2 | intergenic | 0.474 | *NA* | 7 |
| rs36024935 | 14 | 96608042 | C | T | 0.2276 | 0.99 | 2.94E-10 | -0.4675 | 0.0742 | BDKRB2 | intergenic | 0.329 | 5 | 5 |
| rs72704824 | 14 | 96624929 | C | T | 0.2296 | 0.99 | 1.40E-10 | -0.4769 | 0.0743 | BDKRB2 | intergenic | 0.259 | 6 | 5 |
| rs12883511 | 14 | 96617282 | C | T | 0.7664 | 0.97 | 2.62E-10 | 0.4678 | 0.074 | BDKRB2 | intergenic | 0.167 | 7 | 5 |
| rs7144843 | 14 | 96610075 | C | T | 0.7515 | 0.89 | 1.06E-08 | 0.408 | 0.0713 | BDKRB2 | intergenic | 0.146 | 5 | 5 |
| rs112558727 | 14 | 96625519 | C | T | 0.2346 | 0.97 | 1.53E-10 | -0.4753 | 0.0742 | BDKRB2 | intergenic | 0.107 | 6 | 5 |
| rs35526305 | 14 | 96624550 | C | T | 0.2296 | 0.99 | 1.39E-10 | -0.4771 | 0.0743 | BDKRB2 | intergenic | 0.072 | 7 | 5 |
| rs56334881 | 14 | 96608221 | G | A | 0.2276 | 0.99 | 1.44E-10 | -0.4767 | 0.0744 | BDKRB2 | intergenic | 0.004 | 7 | 5 |
| Summary statistics were generated using meta-analysis in METAL. Independent (r^2^ <0.1) lead SNPs in the region are marked red. ANNOVAR = functional variant classification based on position in or outside of a gene; CADD = Combined Annotation-Dependent depletion score, a marker of deleteriousness of SNP's effect on a protein product (higher scores are more deleterious); RDB = RegulomeDB scores, which predict likelihood that a SNP has a regulatory role (lower scores are more likely); minChrState = minimum chromatin state across 127 tissue types (lower scores indicate more open chromatin); commonChrState = most common chromatin state in 127 tissue types; Not Annotated = SNP not available in database. | | | | | | | | | | | | | | |

| **S11 Table.** Expression quantitative trait look-up | | | | | | | | | | | | |
| --- | --- | --- | --- | --- | --- | --- | --- | --- | --- | --- | --- | --- |
| uniqID | db | tissue | gene | testedAllele | p | signed_stats | FDR | RiskIncAllele | alignedDirection | chr | pos | symbol |
| 14:96599503:A:C | GTEx/v8 | Brain_Putamen_basal_ganglia | ENSG00000100739 | A | 3.58E-09 | -0.534 | 5.42E-05 | NA | NA | 14 | 96599503 | BDKRB1 |
| 14:96599503:A:C | GTEx/v8 | Brain_Putamen_basal_ganglia | ENSG00000168398 | A | 3.25E-10 | -0.486 | 1.84E-06 | NA | NA | 14 | 96599503 | BDKRB2 |
| 14:96599656:A:G | GTEx/v8 | Brain_Putamen_basal_ganglia | ENSG00000100739 | A | 3.58E-09 | -0.534 | 5.42E-05 | G | + | 14 | 96599656 | BDKRB1 |
| 14:96599656:A:G | GTEx/v8 | Brain_Putamen_basal_ganglia | ENSG00000168398 | A | 3.25E-10 | -0.486 | 1.84E-06 | G | + | 14 | 96599656 | BDKRB2 |
| 14:96600316:C:T | GTEx/v8 | Brain_Putamen_basal_ganglia | ENSG00000100739 | T | 3.58E-09 | -0.534 | 5.42E-05 | C | + | 14 | 96600316 | BDKRB1 |
| 14:96600316:C:T | GTEx/v8 | Brain_Putamen_basal_ganglia | ENSG00000168398 | T | 3.25E-10 | -0.486 | 1.84E-06 | C | + | 14 | 96600316 | BDKRB2 |
| 14:96600583:C:T | GTEx/v8 | Brain_Putamen_basal_ganglia | ENSG00000100739 | T | 8.25E-09 | -0.521 | 5.42E-05 | C | + | 14 | 96600583 | BDKRB1 |
| 14:96600583:C:T | GTEx/v8 | Brain_Putamen_basal_ganglia | ENSG00000168398 | T | 3.50E-10 | -0.483 | 1.84E-06 | C | + | 14 | 96600583 | BDKRB2 |
| 14:96600596:A:T | GTEx/v8 | Brain_Putamen_basal_ganglia | ENSG00000100739 | A | 3.58E-09 | -0.534 | 5.42E-05 | T | + | 14 | 96600596 | BDKRB1 |
| 14:96600596:A:T | GTEx/v8 | Brain_Putamen_basal_ganglia | ENSG00000168398 | A | 3.25E-10 | -0.486 | 1.84E-06 | T | + | 14 | 96600596 | BDKRB2 |
| 14:96600784:G:T | GTEx/v8 | Brain_Putamen_basal_ganglia | ENSG00000100739 | G | 3.58E-09 | -0.534 | 5.42E-05 | T | + | 14 | 96600784 | BDKRB1 |
| 14:96600784:G:T | GTEx/v8 | Brain_Putamen_basal_ganglia | ENSG00000168398 | G | 3.25E-10 | -0.486 | 1.84E-06 | T | + | 14 | 96600784 | BDKRB2 |
| 14:96602783:A:G | GTEx/v8 | Brain_Putamen_basal_ganglia | ENSG00000100739 | A | 4.97E-09 | 0.513 | 5.42E-05 | A | + | 14 | 96602783 | BDKRB1 |
| 14:96602783:A:G | GTEx/v8 | Brain_Putamen_basal_ganglia | ENSG00000168398 | A | 3.97E-10 | 0.468 | 1.84E-06 | A | + | 14 | 96602783 | BDKRB2 |
| 14:96603267:C:T | GTEx/v8 | Brain_Putamen_basal_ganglia | ENSG00000100739 | T | 4.97E-09 | 0.513 | 5.42E-05 | T | + | 14 | 96603267 | BDKRB1 |
| 14:96603267:C:T | GTEx/v8 | Brain_Putamen_basal_ganglia | ENSG00000168398 | T | 3.97E-10 | 0.468 | 1.84E-06 | T | + | 14 | 96603267 | BDKRB2 |
| 14:96603723:G:T | GTEx/v8 | Brain_Putamen_basal_ganglia | ENSG00000100739 | T | 4.97E-09 | 0.513 | 5.42E-05 | T | + | 14 | 96603723 | BDKRB1 |
| 14:96603723:G:T | GTEx/v8 | Brain_Putamen_basal_ganglia | ENSG00000168398 | T | 3.97E-10 | 0.468 | 1.84E-06 | T | + | 14 | 96603723 | BDKRB2 |
| 14:96603815:C:G | GTEx/v8 | Brain_Putamen_basal_ganglia | ENSG00000100739 | G | 4.97E-09 | 0.513 | 5.42E-05 | G | + | 14 | 96603815 | BDKRB1 |
| 14:96603815:C:G | GTEx/v8 | Brain_Putamen_basal_ganglia | ENSG00000168398 | G | 3.97E-10 | 0.468 | 1.84E-06 | G | + | 14 | 96603815 | BDKRB2 |
| 14:96604435:C:T | GTEx/v8 | Brain_Putamen_basal_ganglia | ENSG00000100739 | T | 4.97E-09 | 0.513 | 5.42E-05 | T | + | 14 | 96604435 | BDKRB1 |
| 14:96604435:C:T | GTEx/v8 | Brain_Putamen_basal_ganglia | ENSG00000168398 | T | 3.97E-10 | 0.468 | 1.84E-06 | T | + | 14 | 96604435 | BDKRB2 |
| 14:96604627:C:T | GTEx/v8 | Brain_Putamen_basal_ganglia | ENSG00000100739 | C | 4.97E-09 | 0.513 | 5.42E-05 | C | + | 14 | 96604627 | BDKRB1 |
| 14:96604627:C:T | GTEx/v8 | Brain_Putamen_basal_ganglia | ENSG00000168398 | C | 3.97E-10 | 0.468 | 1.84E-06 | C | + | 14 | 96604627 | BDKRB2 |
| 14:96605573:A:G | GTEx/v8 | Brain_Putamen_basal_ganglia | ENSG00000100739 | A | 2.66E-08 | -0.509 | 5.42E-05 | G | + | 14 | 96605573 | BDKRB1 |
| 14:96605573:A:G | GTEx/v8 | Brain_Putamen_basal_ganglia | ENSG00000168398 | A | 1.25E-10 | -0.498 | 1.84E-06 | G | + | 14 | 96605573 | BDKRB2 |
| 14:96606733:C:T | GTEx/v8 | Brain_Putamen_basal_ganglia | ENSG00000100739 | T | 5.75E-08 | -0.500 | 5.42E-05 | C | + | 14 | 96606733 | BDKRB1 |
| 14:96606733:C:T | GTEx/v8 | Brain_Putamen_basal_ganglia | ENSG00000168398 | T | 8.85E-11 | -0.503 | 1.84E-06 | C | + | 14 | 96606733 | BDKRB2 |
| 14:96608042:C:T | GTEx/v8 | Brain_Putamen_basal_ganglia | ENSG00000100739 | T | 1.31E-07 | -0.484 | 5.42E-05 | C | + | 14 | 96608042 | BDKRB1 |
| 14:96608042:C:T | GTEx/v8 | Brain_Putamen_basal_ganglia | ENSG00000168398 | T | 1.10E-10 | -0.497 | 1.84E-06 | C | + | 14 | 96608042 | BDKRB2 |
| 14:96608221:A:G | GTEx/v8 | Brain_Putamen_basal_ganglia | ENSG00000100739 | A | 5.75E-08 | -0.500 | 5.42E-05 | G | + | 14 | 96608221 | BDKRB1 |
| 14:96608221:A:G | GTEx/v8 | Brain_Putamen_basal_ganglia | ENSG00000168398 | A | 8.85E-11 | -0.503 | 1.84E-06 | G | + | 14 | 96608221 | BDKRB2 |
| 14:96608962:C:T | GTEx/v8 | Brain_Putamen_basal_ganglia | ENSG00000100739 | T | 1.31E-07 | -0.484 | 5.42E-05 | C | + | 14 | 96608962 | BDKRB1 |
| 14:96608962:C:T | GTEx/v8 | Brain_Putamen_basal_ganglia | ENSG00000168398 | T | 1.10E-10 | -0.497 | 1.84E-06 | C | + | 14 | 96608962 | BDKRB2 |
| 14:96610075:C:T | GTEx/v8 | Brain_Putamen_basal_ganglia | ENSG00000100739 | T | 4.48E-08 | 0.495 | 5.42E-05 | T | + | 14 | 96610075 | BDKRB1 |
| 14:96610075:C:T | GTEx/v8 | Brain_Putamen_basal_ganglia | ENSG00000168398 | T | 1.79E-09 | 0.463 | 1.84E-06 | T | + | 14 | 96610075 | BDKRB2 |
| 14:96611271:C:T | GTEx/v8 | Brain_Putamen_basal_ganglia | ENSG00000100739 | C | 5.75E-08 | -0.500 | 5.42E-05 | T | + | 14 | 96611271 | BDKRB1 |
| 14:96611271:C:T | GTEx/v8 | Brain_Putamen_basal_ganglia | ENSG00000168398 | C | 8.85E-11 | -0.503 | 1.84E-06 | T | + | 14 | 96611271 | BDKRB2 |
| 14:96611391:A:G | GTEx/v8 | Brain_Putamen_basal_ganglia | ENSG00000100739 | G | 5.75E-08 | -0.500 | 5.42E-05 | A | + | 14 | 96611391 | BDKRB1 |
| 14:96611391:A:G | GTEx/v8 | Brain_Putamen_basal_ganglia | ENSG00000168398 | G | 8.85E-11 | -0.503 | 1.84E-06 | A | + | 14 | 96611391 | BDKRB2 |
| 14:96612609:A:G | GTEx/v8 | Brain_Putamen_basal_ganglia | ENSG00000100739 | A | 1.31E-07 | -0.484 | 5.42E-05 | G | + | 14 | 96612609 | BDKRB1 |
| 14:96612609:A:G | GTEx/v8 | Brain_Putamen_basal_ganglia | ENSG00000168398 | A | 1.10E-10 | -0.497 | 1.84E-06 | G | + | 14 | 96612609 | BDKRB2 |
| 14:96612682:C:T | GTEx/v8 | Brain_Putamen_basal_ganglia | ENSG00000100739 | T | 9.73E-08 | 0.482 | 5.42E-05 | T | + | 14 | 96612682 | BDKRB1 |
| 14:96612682:C:T | GTEx/v8 | Brain_Putamen_basal_ganglia | ENSG00000168398 | T | 2.19E-09 | 0.460 | 1.84E-06 | T | + | 14 | 96612682 | BDKRB2 |
| 14:96614325:A:G | GTEx/v8 | Brain_Putamen_basal_ganglia | ENSG00000100739 | G | 4.48E-08 | 0.495 | 5.42E-05 | G | + | 14 | 96614325 | BDKRB1 |
| 14:96614325:A:G | GTEx/v8 | Brain_Putamen_basal_ganglia | ENSG00000168398 | G | 1.79E-09 | 0.463 | 1.84E-06 | G | + | 14 | 96614325 | BDKRB2 |
| 14:96615089:A:G | GTEx/v8 | Brain_Putamen_basal_ganglia | ENSG00000100739 | G | 1.31E-07 | -0.484 | 5.42E-05 | A | + | 14 | 96615089 | BDKRB1 |
| 14:96615089:A:G | GTEx/v8 | Brain_Putamen_basal_ganglia | ENSG00000168398 | G | 1.10E-10 | -0.497 | 1.84E-06 | A | + | 14 | 96615089 | BDKRB2 |
| 14:96615137:A:G | GTEx/v8 | Brain_Putamen_basal_ganglia | ENSG00000100739 | G | 1.31E-07 | -0.484 | 5.42E-05 | A | + | 14 | 96615137 | BDKRB1 |
| 14:96615137:A:G | GTEx/v8 | Brain_Putamen_basal_ganglia | ENSG00000168398 | G | 1.10E-10 | -0.497 | 1.84E-06 | A | + | 14 | 96615137 | BDKRB2 |
| 14:96615980:C:T | GTEx/v8 | Brain_Putamen_basal_ganglia | ENSG00000100739 | C | 1.69E-07 | -0.479 | 5.42E-05 | T | + | 14 | 96615980 | BDKRB1 |
| 14:96615980:C:T | GTEx/v8 | Brain_Putamen_basal_ganglia | ENSG00000168398 | C | 1.93E-09 | -0.466 | 1.84E-06 | T | + | 14 | 96615980 | BDKRB2 |
| 14:96615982:C:T | GTEx/v8 | Brain_Putamen_basal_ganglia | ENSG00000100739 | C | 1.69E-07 | -0.479 | 5.42E-05 | T | + | 14 | 96615982 | BDKRB1 |
| 14:96615982:C:T | GTEx/v8 | Brain_Putamen_basal_ganglia | ENSG00000168398 | C | 1.93E-09 | -0.466 | 1.84E-06 | T | + | 14 | 96615982 | BDKRB2 |
| 14:96616039:C:G | GTEx/v8 | Brain_Putamen_basal_ganglia | ENSG00000100739 | G | 1.69E-07 | -0.479 | 5.42E-05 | C | + | 14 | 96616039 | BDKRB1 |
| 14:96616039:C:G | GTEx/v8 | Brain_Putamen_basal_ganglia | ENSG00000168398 | G | 1.93E-09 | -0.466 | 1.84E-06 | C | + | 14 | 96616039 | BDKRB2 |
| 14:96616330:G:T | GTEx/v8 | Brain_Putamen_basal_ganglia | ENSG00000100739 | T | 9.73E-08 | 0.482 | 5.42E-05 | T | + | 14 | 96616330 | BDKRB1 |
| 14:96616330:G:T | GTEx/v8 | Brain_Putamen_basal_ganglia | ENSG00000168398 | T | 2.19E-09 | 0.460 | 1.84E-06 | T | + | 14 | 96616330 | BDKRB2 |
| 14:96616845:C:T | GTEx/v8 | Brain_Putamen_basal_ganglia | ENSG00000100739 | T | 1.69E-07 | -0.479 | 5.42E-05 | C | + | 14 | 96616845 | BDKRB1 |
| 14:96616845:C:T | GTEx/v8 | Brain_Putamen_basal_ganglia | ENSG00000168398 | T | 1.93E-09 | -0.466 | 1.84E-06 | C | + | 14 | 96616845 | BDKRB2 |
| 14:96617046:A:C | GTEx/v8 | Brain_Putamen_basal_ganglia | ENSG00000100739 | A | 5.75E-08 | -0.500 | 5.42E-05 | C | + | 14 | 96617046 | BDKRB1 |
| 14:96617046:A:C | GTEx/v8 | Brain_Putamen_basal_ganglia | ENSG00000168398 | A | 8.85E-11 | -0.503 | 1.84E-06 | C | + | 14 | 96617046 | BDKRB2 |
| 14:96617205:A:G | GTEx/v8 | Brain_Putamen_basal_ganglia | ENSG00000100739 | A | 1.69E-07 | -0.479 | 5.42E-05 | G | + | 14 | 96617205 | BDKRB1 |
| 14:96617205:A:G | GTEx/v8 | Brain_Putamen_basal_ganglia | ENSG00000168398 | A | 1.93E-09 | -0.466 | 1.84E-06 | G | + | 14 | 96617205 | BDKRB2 |
| 14:96617282:C:T | GTEx/v8 | Brain_Putamen_basal_ganglia | ENSG00000100739 | C | 1.69E-07 | -0.479 | 5.42E-05 | T | + | 14 | 96617282 | BDKRB1 |
| 14:96617282:C:T | GTEx/v8 | Brain_Putamen_basal_ganglia | ENSG00000168398 | C | 1.93E-09 | -0.466 | 1.84E-06 | T | + | 14 | 96617282 | BDKRB2 |
| 14:96617818:A:G | GTEx/v8 | Brain_Putamen_basal_ganglia | ENSG00000100739 | A | 1.69E-07 | -0.479 | 5.42E-05 | G | + | 14 | 96617818 | BDKRB1 |
| 14:96617818:A:G | GTEx/v8 | Brain_Putamen_basal_ganglia | ENSG00000168398 | A | 1.93E-09 | -0.466 | 1.84E-06 | G | + | 14 | 96617818 | BDKRB2 |
| 14:96617826:C:T | GTEx/v8 | Brain_Putamen_basal_ganglia | ENSG00000100739 | T | 1.69E-07 | -0.479 | 5.42E-05 | C | + | 14 | 96617826 | BDKRB1 |
| 14:96617826:C:T | GTEx/v8 | Brain_Putamen_basal_ganglia | ENSG00000168398 | T | 1.93E-09 | -0.466 | 1.84E-06 | C | + | 14 | 96617826 | BDKRB2 |
| 14:96617880:A:G | GTEx/v8 | Brain_Putamen_basal_ganglia | ENSG00000100739 | A | 1.07E-07 | 0.480 | 5.42E-05 | A | + | 14 | 96617880 | BDKRB1 |
| 14:96617880:A:G | GTEx/v8 | Brain_Putamen_basal_ganglia | ENSG00000168398 | A | 2.17E-09 | 0.460 | 1.84E-06 | A | + | 14 | 96617880 | BDKRB2 |
| 14:96617940:C:T | GTEx/v8 | Brain_Putamen_basal_ganglia | ENSG00000100739 | T | 5.75E-08 | -0.500 | 5.42E-05 | C | + | 14 | 96617940 | BDKRB1 |
| 14:96617940:C:T | GTEx/v8 | Brain_Putamen_basal_ganglia | ENSG00000168398 | T | 8.85E-11 | -0.503 | 1.84E-06 | C | + | 14 | 96617940 | BDKRB2 |
| 14:96618312:A:G | GTEx/v8 | Brain_Putamen_basal_ganglia | ENSG00000100739 | A | 1.69E-07 | -0.479 | 5.42E-05 | G | + | 14 | 96618312 | BDKRB1 |
| 14:96618312:A:G | GTEx/v8 | Brain_Putamen_basal_ganglia | ENSG00000168398 | A | 1.93E-09 | -0.466 | 1.84E-06 | G | + | 14 | 96618312 | BDKRB2 |
| 14:96618733:C:G | GTEx/v8 | Brain_Putamen_basal_ganglia | ENSG00000100739 | G | 1.69E-07 | -0.479 | 5.42E-05 | C | + | 14 | 96618733 | BDKRB1 |
| 14:96618733:C:G | GTEx/v8 | Brain_Putamen_basal_ganglia | ENSG00000168398 | G | 1.93E-09 | -0.466 | 1.84E-06 | C | + | 14 | 96618733 | BDKRB2 |
| 14:96618740:A:C | GTEx/v8 | Brain_Putamen_basal_ganglia | ENSG00000100739 | C | 1.69E-07 | -0.479 | 5.42E-05 | A | + | 14 | 96618740 | BDKRB1 |
| 14:96618740:A:C | GTEx/v8 | Brain_Putamen_basal_ganglia | ENSG00000168398 | C | 1.93E-09 | -0.466 | 1.84E-06 | A | + | 14 | 96618740 | BDKRB2 |
| 14:96618781:A:T | GTEx/v8 | Brain_Putamen_basal_ganglia | ENSG00000100739 | T | 1.69E-07 | -0.479 | 5.42E-05 | A | + | 14 | 96618781 | BDKRB1 |
| 14:96618781:A:T | GTEx/v8 | Brain_Putamen_basal_ganglia | ENSG00000168398 | T | 1.93E-09 | -0.466 | 1.84E-06 | A | + | 14 | 96618781 | BDKRB2 |
| 14:96619307:G:T | GTEx/v8 | Brain_Putamen_basal_ganglia | ENSG00000100739 | T | 1.69E-07 | -0.479 | 5.42E-05 | G | + | 14 | 96619307 | BDKRB1 |
| 14:96619307:G:T | GTEx/v8 | Brain_Putamen_basal_ganglia | ENSG00000168398 | T | 1.93E-09 | -0.466 | 1.84E-06 | G | + | 14 | 96619307 | BDKRB2 |
| 14:96619480:A:G | GTEx/v8 | Brain_Putamen_basal_ganglia | ENSG00000100739 | G | 1.69E-07 | -0.479 | 5.42E-05 | A | + | 14 | 96619480 | BDKRB1 |
| 14:96619480:A:G | GTEx/v8 | Brain_Putamen_basal_ganglia | ENSG00000168398 | G | 1.93E-09 | -0.466 | 1.84E-06 | A | + | 14 | 96619480 | BDKRB2 |
| 14:96619739:G:T | GTEx/v8 | Brain_Putamen_basal_ganglia | ENSG00000100739 | T | 1.69E-07 | -0.479 | 5.42E-05 | G | + | 14 | 96619739 | BDKRB1 |
| 14:96619739:G:T | GTEx/v8 | Brain_Putamen_basal_ganglia | ENSG00000168398 | T | 1.93E-09 | -0.466 | 1.84E-06 | G | + | 14 | 96619739 | BDKRB2 |
| 14:96620441:A:G | GTEx/v8 | Brain_Putamen_basal_ganglia | ENSG00000100739 | G | 1.69E-07 | -0.479 | 5.42E-05 | A | + | 14 | 96620441 | BDKRB1 |
| 14:96620441:A:G | GTEx/v8 | Brain_Putamen_basal_ganglia | ENSG00000168398 | G | 1.93E-09 | -0.466 | 1.84E-06 | A | + | 14 | 96620441 | BDKRB2 |
| 14:96620508:A:G | GTEx/v8 | Brain_Putamen_basal_ganglia | ENSG00000100739 | A | 1.69E-07 | -0.479 | 5.42E-05 | G | + | 14 | 96620508 | BDKRB1 |
| 14:96620508:A:G | GTEx/v8 | Brain_Putamen_basal_ganglia | ENSG00000168398 | A | 1.93E-09 | -0.466 | 1.84E-06 | G | + | 14 | 96620508 | BDKRB2 |
| 14:96620556:A:G | GTEx/v8 | Brain_Putamen_basal_ganglia | ENSG00000100739 | G | 1.69E-07 | -0.479 | 5.42E-05 | A | + | 14 | 96620556 | BDKRB1 |
| 14:96620556:A:G | GTEx/v8 | Brain_Putamen_basal_ganglia | ENSG00000168398 | G | 1.93E-09 | -0.466 | 1.84E-06 | A | + | 14 | 96620556 | BDKRB2 |
| 14:96620639:A:G | GTEx/v8 | Brain_Putamen_basal_ganglia | ENSG00000100739 | G | 1.69E-07 | -0.479 | 5.42E-05 | A | + | 14 | 96620639 | BDKRB1 |
| 14:96620639:A:G | GTEx/v8 | Brain_Putamen_basal_ganglia | ENSG00000168398 | G | 1.93E-09 | -0.466 | 1.84E-06 | A | + | 14 | 96620639 | BDKRB2 |
| 14:96620826:A:G | GTEx/v8 | Brain_Putamen_basal_ganglia | ENSG00000100739 | G | 1.97E-07 | -0.475 | 5.42E-05 | A | + | 14 | 96620826 | BDKRB1 |
| 14:96620826:A:G | GTEx/v8 | Brain_Putamen_basal_ganglia | ENSG00000168398 | G | 1.64E-09 | -0.467 | 1.84E-06 | A | + | 14 | 96620826 | BDKRB2 |
| 14:96620874:C:T | GTEx/v8 | Brain_Putamen_basal_ganglia | ENSG00000100739 | T | 1.69E-07 | -0.479 | 5.42E-05 | C | + | 14 | 96620874 | BDKRB1 |
| 14:96620874:C:T | GTEx/v8 | Brain_Putamen_basal_ganglia | ENSG00000168398 | T | 1.93E-09 | -0.466 | 1.84E-06 | C | + | 14 | 96620874 | BDKRB2 |
| 14:96620955:A:G | GTEx/v8 | Brain_Putamen_basal_ganglia | ENSG00000100739 | G | 1.69E-07 | -0.479 | 5.42E-05 | A | + | 14 | 96620955 | BDKRB1 |
| 14:96620955:A:G | GTEx/v8 | Brain_Putamen_basal_ganglia | ENSG00000168398 | G | 1.93E-09 | -0.466 | 1.84E-06 | A | + | 14 | 96620955 | BDKRB2 |
| 14:96621379:A:G | GTEx/v8 | Brain_Putamen_basal_ganglia | ENSG00000100739 | G | 1.69E-07 | -0.479 | 5.42E-05 | A | + | 14 | 96621379 | BDKRB1 |
| 14:96621379:A:G | GTEx/v8 | Brain_Putamen_basal_ganglia | ENSG00000168398 | G | 1.93E-09 | -0.466 | 1.84E-06 | A | + | 14 | 96621379 | BDKRB2 |
| 14:96621809:A:G | GTEx/v8 | Brain_Putamen_basal_ganglia | ENSG00000100739 | A | 1.69E-07 | -0.479 | 5.42E-05 | G | + | 14 | 96621809 | BDKRB1 |
| 14:96621809:A:G | GTEx/v8 | Brain_Putamen_basal_ganglia | ENSG00000168398 | A | 1.93E-09 | -0.466 | 1.84E-06 | G | + | 14 | 96621809 | BDKRB2 |
| 14:96621833:C:G | GTEx/v8 | Brain_Putamen_basal_ganglia | ENSG00000100739 | G | 1.69E-07 | -0.479 | 5.42E-05 | C | + | 14 | 96621833 | BDKRB1 |
| 14:96621833:C:G | GTEx/v8 | Brain_Putamen_basal_ganglia | ENSG00000168398 | G | 1.93E-09 | -0.466 | 1.84E-06 | C | + | 14 | 96621833 | BDKRB2 |
| 14:96622427:C:T | GTEx/v8 | Brain_Putamen_basal_ganglia | ENSG00000100739 | C | 1.07E-07 | 0.480 | 5.42E-05 | C | + | 14 | 96622427 | BDKRB1 |
| 14:96622427:C:T | GTEx/v8 | Brain_Putamen_basal_ganglia | ENSG00000168398 | C | 2.17E-09 | 0.460 | 1.84E-06 | C | + | 14 | 96622427 | BDKRB2 |
| 14:96622909:A:C | GTEx/v8 | Brain_Putamen_basal_ganglia | ENSG00000100739 | C | 2.20E-07 | 0.469 | 5.42E-05 | C | + | 14 | 96622909 | BDKRB1 |
| 14:96622909:A:C | GTEx/v8 | Brain_Putamen_basal_ganglia | ENSG00000168398 | C | 2.58E-09 | 0.457 | 1.84E-06 | C | + | 14 | 96622909 | BDKRB2 |
| 14:96624517:A:G | GTEx/v8 | Brain_Putamen_basal_ganglia | ENSG00000100739 | G | 2.33E-07 | -0.472 | 5.42E-05 | A | + | 14 | 96624517 | BDKRB1 |
| 14:96624517:A:G | GTEx/v8 | Brain_Putamen_basal_ganglia | ENSG00000168398 | G | 2.70E-10 | -0.484 | 1.84E-06 | A | + | 14 | 96624517 | BDKRB2 |
| 14:96624550:C:T | GTEx/v8 | Brain_Putamen_basal_ganglia | ENSG00000100739 | T | 2.33E-07 | -0.472 | 5.42E-05 | C | + | 14 | 96624550 | BDKRB1 |
| 14:96624550:C:T | GTEx/v8 | Brain_Putamen_basal_ganglia | ENSG00000168398 | T | 2.70E-10 | -0.484 | 1.84E-06 | C | + | 14 | 96624550 | BDKRB2 |
| 14:96624719:G:T | GTEx/v8 | Brain_Putamen_basal_ganglia | ENSG00000100739 | T | 2.33E-07 | -0.472 | 5.42E-05 | G | + | 14 | 96624719 | BDKRB1 |
| 14:96624719:G:T | GTEx/v8 | Brain_Putamen_basal_ganglia | ENSG00000168398 | T | 2.70E-10 | -0.484 | 1.84E-06 | G | + | 14 | 96624719 | BDKRB2 |
| 14:96624929:C:T | GTEx/v8 | Brain_Putamen_basal_ganglia | ENSG00000100739 | T | 7.65E-08 | -0.488 | 5.42E-05 | C | + | 14 | 96624929 | BDKRB1 |
| 14:96624929:C:T | GTEx/v8 | Brain_Putamen_basal_ganglia | ENSG00000168398 | T | 4.10E-10 | -0.480 | 1.84E-06 | C | + | 14 | 96624929 | BDKRB2 |
| 14:96625519:C:T | GTEx/v8 | Brain_Putamen_basal_ganglia | ENSG00000100739 | T | 3.04E-07 | -0.472 | 5.42E-05 | C | + | 14 | 96625519 | BDKRB1 |
| 14:96625519:C:T | GTEx/v8 | Brain_Putamen_basal_ganglia | ENSG00000168398 | T | 5.22E-09 | -0.457 | 1.84E-06 | C | + | 14 | 96625519 | BDKRB2 |
| 14:96625772:C:T | GTEx/v8 | Brain_Putamen_basal_ganglia | ENSG00000100739 | C | 3.04E-07 | -0.472 | 5.42E-05 | T | + | 14 | 96625772 | BDKRB1 |
| 14:96625772:C:T | GTEx/v8 | Brain_Putamen_basal_ganglia | ENSG00000168398 | C | 5.22E-09 | -0.457 | 1.84E-06 | T | + | 14 | 96625772 | BDKRB2 |
| 14:96599503:A:C | GTEx/v8 | Lung | ENSG00000168398 | A | 1.13E-05 | 0.188 | 2.96E-13 | NA | NA | 14 | 96599503 | BDKRB2 |
| 14:96599656:A:G | GTEx/v8 | Lung | ENSG00000168398 | A | 1.15E-05 | 0.188 | 2.96E-13 | G | - | 14 | 96599656 | BDKRB2 |
| 14:96600316:C:T | GTEx/v8 | Lung | ENSG00000168398 | T | 6.63E-06 | 0.192 | 2.96E-13 | C | - | 14 | 96600316 | BDKRB2 |
| 14:96600583:C:T | GTEx/v8 | Lung | ENSG00000168398 | T | 5.09E-06 | 0.195 | 2.96E-13 | C | - | 14 | 96600583 | BDKRB2 |
| 14:96600596:A:T | GTEx/v8 | Lung | ENSG00000168398 | A | 6.63E-06 | 0.192 | 2.96E-13 | T | - | 14 | 96600596 | BDKRB2 |
| 14:96600784:G:T | GTEx/v8 | Lung | ENSG00000168398 | G | 6.63E-06 | 0.192 | 2.96E-13 | T | - | 14 | 96600784 | BDKRB2 |
| 14:96602783:A:G | GTEx/v8 | Lung | ENSG00000168398 | A | 1.57E-07 | -0.215 | 2.96E-13 | A | - | 14 | 96602783 | BDKRB2 |
| 14:96603267:C:T | GTEx/v8 | Lung | ENSG00000168398 | T | 1.57E-07 | -0.215 | 2.96E-13 | T | - | 14 | 96603267 | BDKRB2 |
| 14:96603723:G:T | GTEx/v8 | Lung | ENSG00000168398 | T | 1.57E-07 | -0.215 | 2.96E-13 | T | - | 14 | 96603723 | BDKRB2 |
| 14:96603815:C:G | GTEx/v8 | Lung | ENSG00000168398 | G | 1.57E-07 | -0.215 | 2.96E-13 | G | - | 14 | 96603815 | BDKRB2 |
| 14:96604435:C:T | GTEx/v8 | Lung | ENSG00000168398 | T | 1.57E-07 | -0.215 | 2.96E-13 | T | - | 14 | 96604435 | BDKRB2 |
| 14:96604627:C:T | GTEx/v8 | Lung | ENSG00000168398 | C | 1.57E-07 | -0.215 | 2.96E-13 | C | - | 14 | 96604627 | BDKRB2 |
| 14:96605573:A:G | GTEx/v8 | Lung | ENSG00000168398 | A | 3.25E-06 | 0.202 | 2.96E-13 | G | - | 14 | 96605573 | BDKRB2 |
| 14:96606733:C:T | GTEx/v8 | Lung | ENSG00000168398 | T | 2.45E-06 | 0.202 | 2.96E-13 | C | - | 14 | 96606733 | BDKRB2 |
| 14:96608042:C:T | GTEx/v8 | Lung | ENSG00000168398 | T | 2.99E-06 | 0.201 | 2.96E-13 | C | - | 14 | 96608042 | BDKRB2 |
| 14:96608221:A:G | GTEx/v8 | Lung | ENSG00000168398 | A | 3.88E-06 | 0.198 | 2.96E-13 | G | - | 14 | 96608221 | BDKRB2 |
| 14:96608962:C:T | GTEx/v8 | Lung | ENSG00000168398 | T | 3.89E-06 | 0.198 | 2.96E-13 | C | - | 14 | 96608962 | BDKRB2 |
| 14:96610075:C:T | GTEx/v8 | Lung | ENSG00000168398 | T | 1.64E-08 | -0.230 | 2.96E-13 | T | - | 14 | 96610075 | BDKRB2 |
| 14:96611271:C:T | GTEx/v8 | Lung | ENSG00000168398 | C | 1.68E-06 | 0.205 | 2.96E-13 | T | - | 14 | 96611271 | BDKRB2 |
| 14:96611391:A:G | GTEx/v8 | Lung | ENSG00000168398 | G | 2.05E-06 | 0.203 | 2.96E-13 | A | - | 14 | 96611391 | BDKRB2 |
| 14:96612609:A:G | GTEx/v8 | Lung | ENSG00000168398 | A | 1.58E-06 | 0.205 | 2.96E-13 | G | - | 14 | 96612609 | BDKRB2 |
| 14:96612682:C:T | GTEx/v8 | Lung | ENSG00000168398 | T | 2.36E-08 | -0.228 | 2.96E-13 | T | - | 14 | 96612682 | BDKRB2 |
| 14:96614325:A:G | GTEx/v8 | Lung | ENSG00000168398 | G | 2.18E-08 | -0.227 | 2.96E-13 | G | - | 14 | 96614325 | BDKRB2 |
| 14:96615089:A:G | GTEx/v8 | Lung | ENSG00000168398 | G | 1.30E-06 | 0.207 | 2.96E-13 | A | - | 14 | 96615089 | BDKRB2 |
| 14:96615137:A:G | GTEx/v8 | Lung | ENSG00000168398 | G | 1.30E-06 | 0.207 | 2.96E-13 | A | - | 14 | 96615137 | BDKRB2 |
| 14:96615980:C:T | GTEx/v8 | Lung | ENSG00000168398 | C | 2.54E-06 | 0.196 | 2.96E-13 | T | - | 14 | 96615980 | BDKRB2 |
| 14:96615982:C:T | GTEx/v8 | Lung | ENSG00000168398 | C | 2.54E-06 | 0.196 | 2.96E-13 | T | - | 14 | 96615982 | BDKRB2 |
| 14:96616039:C:G | GTEx/v8 | Lung | ENSG00000168398 | G | 2.54E-06 | 0.196 | 2.96E-13 | C | - | 14 | 96616039 | BDKRB2 |
| 14:96616330:G:T | GTEx/v8 | Lung | ENSG00000168398 | T | 1.52E-08 | -0.230 | 2.96E-13 | T | - | 14 | 96616330 | BDKRB2 |
| 14:96616845:C:T | GTEx/v8 | Lung | ENSG00000168398 | T | 2.58E-06 | 0.196 | 2.96E-13 | C | - | 14 | 96616845 | BDKRB2 |
| 14:96617046:A:C | GTEx/v8 | Lung | ENSG00000168398 | A | 1.27E-06 | 0.207 | 2.96E-13 | C | - | 14 | 96617046 | BDKRB2 |
| 14:96617205:A:G | GTEx/v8 | Lung | ENSG00000168398 | A | 1.96E-06 | 0.198 | 2.96E-13 | G | - | 14 | 96617205 | BDKRB2 |
| 14:96617282:C:T | GTEx/v8 | Lung | ENSG00000168398 | C | 2.54E-06 | 0.196 | 2.96E-13 | T | - | 14 | 96617282 | BDKRB2 |
| 14:96617818:A:G | GTEx/v8 | Lung | ENSG00000168398 | A | 2.54E-06 | 0.196 | 2.96E-13 | G | - | 14 | 96617818 | BDKRB2 |
| 14:96617826:C:T | GTEx/v8 | Lung | ENSG00000168398 | T | 2.54E-06 | 0.196 | 2.96E-13 | C | - | 14 | 96617826 | BDKRB2 |
| 14:96617880:A:G | GTEx/v8 | Lung | ENSG00000168398 | A | 2.42E-08 | -0.226 | 2.96E-13 | A | - | 14 | 96617880 | BDKRB2 |
| 14:96617940:C:T | GTEx/v8 | Lung | ENSG00000168398 | T | 1.27E-06 | 0.207 | 2.96E-13 | C | - | 14 | 96617940 | BDKRB2 |
| 14:96618312:A:G | GTEx/v8 | Lung | ENSG00000168398 | A | 2.54E-06 | 0.196 | 2.96E-13 | G | - | 14 | 96618312 | BDKRB2 |
| 14:96618733:C:G | GTEx/v8 | Lung | ENSG00000168398 | G | 2.54E-06 | 0.196 | 2.96E-13 | C | - | 14 | 96618733 | BDKRB2 |
| 14:96618740:A:C | GTEx/v8 | Lung | ENSG00000168398 | C | 2.54E-06 | 0.196 | 2.96E-13 | A | - | 14 | 96618740 | BDKRB2 |
| 14:96618781:A:T | GTEx/v8 | Lung | ENSG00000168398 | T | 2.54E-06 | 0.196 | 2.96E-13 | A | - | 14 | 96618781 | BDKRB2 |
| 14:96619307:G:T | GTEx/v8 | Lung | ENSG00000168398 | T | 1.43E-06 | 0.201 | 2.96E-13 | G | - | 14 | 96619307 | BDKRB2 |
| 14:96619480:A:G | GTEx/v8 | Lung | ENSG00000168398 | G | 1.43E-06 | 0.201 | 2.96E-13 | A | - | 14 | 96619480 | BDKRB2 |
| 14:96619739:G:T | GTEx/v8 | Lung | ENSG00000168398 | T | 1.43E-06 | 0.201 | 2.96E-13 | G | - | 14 | 96619739 | BDKRB2 |
| 14:96620441:A:G | GTEx/v8 | Lung | ENSG00000168398 | G | 1.43E-06 | 0.201 | 2.96E-13 | A | - | 14 | 96620441 | BDKRB2 |
| 14:96620508:A:G | GTEx/v8 | Lung | ENSG00000168398 | A | 1.43E-06 | 0.201 | 2.96E-13 | G | - | 14 | 96620508 | BDKRB2 |
| 14:96620556:A:G | GTEx/v8 | Lung | ENSG00000168398 | G | 1.43E-06 | 0.201 | 2.96E-13 | A | - | 14 | 96620556 | BDKRB2 |
| 14:96620639:A:G | GTEx/v8 | Lung | ENSG00000168398 | G | 1.43E-06 | 0.201 | 2.96E-13 | A | - | 14 | 96620639 | BDKRB2 |
| 14:96620826:A:G | GTEx/v8 | Lung | ENSG00000168398 | G | 1.45E-06 | 0.201 | 2.96E-13 | A | - | 14 | 96620826 | BDKRB2 |
| 14:96620874:C:T | GTEx/v8 | Lung | ENSG00000168398 | T | 1.43E-06 | 0.201 | 2.96E-13 | C | - | 14 | 96620874 | BDKRB2 |
| 14:96620955:A:G | GTEx/v8 | Lung | ENSG00000168398 | G | 1.43E-06 | 0.201 | 2.96E-13 | A | - | 14 | 96620955 | BDKRB2 |
| 14:96621379:A:G | GTEx/v8 | Lung | ENSG00000168398 | G | 1.43E-06 | 0.201 | 2.96E-13 | A | - | 14 | 96621379 | BDKRB2 |
| 14:96621809:A:G | GTEx/v8 | Lung | ENSG00000168398 | A | 1.09E-06 | 0.203 | 2.96E-13 | G | - | 14 | 96621809 | BDKRB2 |
| 14:96621833:C:G | GTEx/v8 | Lung | ENSG00000168398 | G | 1.43E-06 | 0.201 | 2.96E-13 | C | - | 14 | 96621833 | BDKRB2 |
| 14:96622427:C:T | GTEx/v8 | Lung | ENSG00000168398 | C | 1.33E-08 | -0.229 | 2.96E-13 | C | - | 14 | 96622427 | BDKRB2 |
| 14:96622909:A:C | GTEx/v8 | Lung | ENSG00000168398 | C | 1.60E-08 | -0.230 | 2.96E-13 | C | - | 14 | 96622909 | BDKRB2 |
| 14:96624517:A:G | GTEx/v8 | Lung | ENSG00000168398 | G | 2.24E-06 | 0.200 | 2.96E-13 | A | - | 14 | 96624517 | BDKRB2 |
| 14:96624550:C:T | GTEx/v8 | Lung | ENSG00000168398 | T | 2.24E-06 | 0.200 | 2.96E-13 | C | - | 14 | 96624550 | BDKRB2 |
| 14:96624719:G:T | GTEx/v8 | Lung | ENSG00000168398 | T | 1.17E-06 | 0.206 | 2.96E-13 | G | - | 14 | 96624719 | BDKRB2 |
| 14:96624929:C:T | GTEx/v8 | Lung | ENSG00000168398 | T | 1.91E-06 | 0.200 | 2.96E-13 | C | - | 14 | 96624929 | BDKRB2 |
| 14:96625519:C:T | GTEx/v8 | Lung | ENSG00000168398 | T | 3.22E-06 | 0.196 | 2.96E-13 | C | - | 14 | 96625519 | BDKRB2 |
| 14:96625772:C:T | GTEx/v8 | Lung | ENSG00000168398 | C | 3.96E-06 | 0.195 | 2.96E-13 | T | - | 14 | 96625772 | BDKRB2 |

| **S12 Table.** Genes implicated by positional, eQTL, or chromatin interaction mapping of SNPs associated with ACE-inhibitor associated angioedema | | | | | | | | | | | | | | | | | | |
| --- | --- | --- | --- | --- | --- | --- | --- | --- | --- | --- | --- | --- | --- | --- | --- | --- | --- | --- |
| **Gene** | **Ensembl ID** | **CHR** | **Start BP** | **End BP** | **pLI** | **posMap #SNPs** | **posMap Max CADD** | **eqtlMap #SNPs** | **eqtlMap minP** | **eqtlMap minQ** | **eqtlMap Tissues** | **eqtlMap Direction** | **ciMap** | **ciMap Tissues** | **coloc PP4** | **coloc Tissues** | **minGwasP** | **IndSigSNPs** |
| RP11-1070N10.3 | ENSG00000258572 | 14 | 95982473 | 95984248 | *NA* | 0 | 0 | 0 | *NA* | *NA* | *NA* | *NA* | Yes | IMR90:Mesenchymal_Stem_Cell | - |  | 1.62E-10 | rs12888576 |
| GLRX5 | ENSG00000182512 | 14 | 95999840 | 96011061 | 0.6262 | 0 | 0 | 0 | *NA* | *NA* | *NA* | *NA* | Yes | Left_Ventricle:IMR90:Mesenchymal_Stem_Cell:Mesendoderm:Trophoblast-like_Cell:hESC | - |  | 1.62E-10 | rs12888576 |
| TCL1A | ENSG00000100721 | 14 | 96176304 | 96180533 | 0.0012 | 0 | 0 | 0 | *NA* | *NA* | *NA* | *NA* | Yes | hESC | - |  | 1.62E-10 | rs12888576 |
| C14orf132 | ENSG00000227051 | 14 | 96505661 | 96560417 | *NA* | 67 | 18.11 | 0 | *NA* | *NA* | *NA* | *NA* | No | *NA* | 0.00 | gtex_v8_brain_putamen_basal_ganglia | 1.02E-10 | rs12888576 |
| BDKRB2 | ENSG00000168398 | 14 | 96671016 | 96710666 | 0.2699 | 67 | 18.11 | 62 | 8.85E-11 | 2.96E-13 | GTEx/v8/Brain_Putamen_basal_ganglia:GTEx/v8/Lung | - | Yes | IMR90 | 0.95 | gtex_v8_brain_putamen_basal_ganglia | 1.02E-10 | rs12888576 |
| RP11-404P21.8 | ENSG00000258691 | 14 | 96671181 | 96730266 | *NA* | 67 | 18.11 | 0 | *NA* | *NA* | *NA* | *NA* | Yes | IMR90 | - |  | 1.02E-10 | rs12888576 |
| BDKRB1 | ENSG00000100739 | 14 | 96722161 | 96735304 | 0.0701 | 10 | 5.101 | 62 | 3.58E-09 | 5.42E-05 | GTEx/v8/Brain_Putamen_basal_ganglia | + | No | *NA* | 0.90 | gtex_v8_brain_putamen_basal_ganglia | 1.02E-10 | rs12888576 |
| SNPs were identified by a genome-wide significant (Bonferroni-corrected P<5e-8) meta-analysis and mapped to genes using positional mapping, eQTL mapping, and chromatin interaction strategies. pLI = probability of loss of function mutation intolerance (higher scores indicate higher probability); posMap = SNPs mapped to this gene by positional mapping; CADD = Combined Annotation-Dependent depletion score (higher scores indicate more deleterious effects of a SNP); eqtlMap = SNPs mapped to this gene by expression quantitative traic locus mapping; minP = minimum P value of eQTL association for the mapped gene; minQ = minimum false discovery rate Q value of eQTL association for the mapped gene; Tissues = tissues in which the significant eQTL association or chromatin interaction was observed; Direction = direction of association of eQTL SNP with expression levels (upregulation or downregulation); ci = chromatin interaction mapping; minGwasP = minimum GWAS P value of SNP(s) implicating the gene; IndSigSNPs = SNPs with significant GWAS p-values from which the gene was mapped. | | | | | | | | | | | | | | | | | | |

| **S13 Table.** Chromatin interaction regions linking significant GWAS loci from to implicated genes. | | | | | |
| --- | --- | --- | --- | --- | --- |
| **InteractionRegion 1** | **InteractionRegion 2** | **FDR** | **Tissue/Cell** | **GWS SNPs in Interaction Region** | **Genes in Interaction Region** |
| 14:96600001-96640000 | 14:96000001-96040000 | 1.83E-09 | Left_Ventricle | rs59804216;rs2369539;rs4627266;rs2369541;rs4905447;rs7156430;rs1889374;rs1889372;rs11160314;rs3939400;rs55940712;rs60634508;rs36024935;rs56334881;rs2151767;rs7144843;rs34485356;rs12888576;rs72704813;rs12894970;rs369235957;rs12885218;rs71415026;rs68023675;rs35262593;rs34857325;rs36092996;rs4905449;rs10140368;rs8022837;rs34845487;rs35974883;rs12883511;rs2369542;rs8008696;rs8008421;rs1959041;rs34985854;rs11850248;rs11850332;rs11850303;rs11850334;rs55668608;rs35136400;rs12894873;rs11846378;rs11846531;rs11846417;rs11846465;rs11846550;rs71415027;rs71415028;rs2369544;rs28849215;rs28823359;rs12432014;rs61985296;rs12881275;rs7492727;rs34393530;rs35526305;rs34870532;rs72704824;rs112558727;rs66680728 | ENSG00000182512 |
| 14:96600001-96640000 | 14:96640001-96680000 | 6.55E-07 | IMR90 | rs59804216;rs2369539;rs4627266;rs2369541;rs4905447;rs7156430;rs1889374;rs1889372;rs11160314;rs3939400;rs55940712;rs60634508;rs36024935;rs56334881;rs2151767;rs7144843;rs34485356;rs12888576;rs72704813;rs12894970;rs369235957;rs12885218;rs71415026;rs68023675;rs35262593;rs34857325;rs36092996;rs4905449;rs10140368;rs8022837;rs34845487;rs35974883;rs12883511;rs2369542;rs8008696;rs8008421;rs1959041;rs34985854;rs11850248;rs11850332;rs11850303;rs11850334;rs55668608;rs35136400;rs12894873;rs11846378;rs11846531;rs11846417;rs11846465;rs11846550;rs71415027;rs71415028;rs2369544;rs28849215;rs28823359;rs12432014;rs61985296;rs12881275;rs7492727;rs34393530;rs35526305;rs34870532;rs72704824;rs112558727;rs66680728 | ENSG00000168398:ENSG00000258691 |
| 14:96600001-96640000 | 14:95960001-96000000 | 2.86E-10 | IMR90 | rs59804216;rs2369539;rs4627266;rs2369541;rs4905447;rs7156430;rs1889374;rs1889372;rs11160314;rs3939400;rs55940712;rs60634508;rs36024935;rs56334881;rs2151767;rs7144843;rs34485356;rs12888576;rs72704813;rs12894970;rs369235957;rs12885218;rs71415026;rs68023675;rs35262593;rs34857325;rs36092996;rs4905449;rs10140368;rs8022837;rs34845487;rs35974883;rs12883511;rs2369542;rs8008696;rs8008421;rs1959041;rs34985854;rs11850248;rs11850332;rs11850303;rs11850334;rs55668608;rs35136400;rs12894873;rs11846378;rs11846531;rs11846417;rs11846465;rs11846550;rs71415027;rs71415028;rs2369544;rs28849215;rs28823359;rs12432014;rs61985296;rs12881275;rs7492727;rs34393530;rs35526305;rs34870532;rs72704824;rs112558727;rs66680728 | ENSG00000258572:ENSG00000182512 |
| 14:96600001-96640000 | 14:96000001-96040000 | 3.37E-27 | IMR90 | rs59804216;rs2369539;rs4627266;rs2369541;rs4905447;rs7156430;rs1889374;rs1889372;rs11160314;rs3939400;rs55940712;rs60634508;rs36024935;rs56334881;rs2151767;rs7144843;rs34485356;rs12888576;rs72704813;rs12894970;rs369235957;rs12885218;rs71415026;rs68023675;rs35262593;rs34857325;rs36092996;rs4905449;rs10140368;rs8022837;rs34845487;rs35974883;rs12883511;rs2369542;rs8008696;rs8008421;rs1959041;rs34985854;rs11850248;rs11850332;rs11850303;rs11850334;rs55668608;rs35136400;rs12894873;rs11846378;rs11846531;rs11846417;rs11846465;rs11846550;rs71415027;rs71415028;rs2369544;rs28849215;rs28823359;rs12432014;rs61985296;rs12881275;rs7492727;rs34393530;rs35526305;rs34870532;rs72704824;rs112558727;rs66680728 | ENSG00000182512 |
| 14:96600001-96640000 | 14:95960001-96000000 | 2.14E-08 | Mesenchymal_Stem_Cell | rs59804216;rs2369539;rs4627266;rs2369541;rs4905447;rs7156430;rs1889374;rs1889372;rs11160314;rs3939400;rs55940712;rs60634508;rs36024935;rs56334881;rs2151767;rs7144843;rs34485356;rs12888576;rs72704813;rs12894970;rs369235957;rs12885218;rs71415026;rs68023675;rs35262593;rs34857325;rs36092996;rs4905449;rs10140368;rs8022837;rs34845487;rs35974883;rs12883511;rs2369542;rs8008696;rs8008421;rs1959041;rs34985854;rs11850248;rs11850332;rs11850303;rs11850334;rs55668608;rs35136400;rs12894873;rs11846378;rs11846531;rs11846417;rs11846465;rs11846550;rs71415027;rs71415028;rs2369544;rs28849215;rs28823359;rs12432014;rs61985296;rs12881275;rs7492727;rs34393530;rs35526305;rs34870532;rs72704824;rs112558727;rs66680728 | ENSG00000258572:ENSG00000182512 |
| 14:96600001-96640000 | 14:96000001-96040000 | 7.65E-45 | Mesenchymal_Stem_Cell | rs59804216;rs2369539;rs4627266;rs2369541;rs4905447;rs7156430;rs1889374;rs1889372;rs11160314;rs3939400;rs55940712;rs60634508;rs36024935;rs56334881;rs2151767;rs7144843;rs34485356;rs12888576;rs72704813;rs12894970;rs369235957;rs12885218;rs71415026;rs68023675;rs35262593;rs34857325;rs36092996;rs4905449;rs10140368;rs8022837;rs34845487;rs35974883;rs12883511;rs2369542;rs8008696;rs8008421;rs1959041;rs34985854;rs11850248;rs11850332;rs11850303;rs11850334;rs55668608;rs35136400;rs12894873;rs11846378;rs11846531;rs11846417;rs11846465;rs11846550;rs71415027;rs71415028;rs2369544;rs28849215;rs28823359;rs12432014;rs61985296;rs12881275;rs7492727;rs34393530;rs35526305;rs34870532;rs72704824;rs112558727;rs66680728 | ENSG00000182512 |
| 14:96600001-96640000 | 14:96000001-96040000 | 3.70E-52 | Mesendoderm | rs59804216;rs2369539;rs4627266;rs2369541;rs4905447;rs7156430;rs1889374;rs1889372;rs11160314;rs3939400;rs55940712;rs60634508;rs36024935;rs56334881;rs2151767;rs7144843;rs34485356;rs12888576;rs72704813;rs12894970;rs369235957;rs12885218;rs71415026;rs68023675;rs35262593;rs34857325;rs36092996;rs4905449;rs10140368;rs8022837;rs34845487;rs35974883;rs12883511;rs2369542;rs8008696;rs8008421;rs1959041;rs34985854;rs11850248;rs11850332;rs11850303;rs11850334;rs55668608;rs35136400;rs12894873;rs11846378;rs11846531;rs11846417;rs11846465;rs11846550;rs71415027;rs71415028;rs2369544;rs28849215;rs28823359;rs12432014;rs61985296;rs12881275;rs7492727;rs34393530;rs35526305;rs34870532;rs72704824;rs112558727;rs66680728 | ENSG00000182512 |
| 14:96600001-96640000 | 14:96000001-96040000 | 7.36E-14 | Trophoblast-like_Cell | rs59804216;rs2369539;rs4627266;rs2369541;rs4905447;rs7156430;rs1889374;rs1889372;rs11160314;rs3939400;rs55940712;rs60634508;rs36024935;rs56334881;rs2151767;rs7144843;rs34485356;rs12888576;rs72704813;rs12894970;rs369235957;rs12885218;rs71415026;rs68023675;rs35262593;rs34857325;rs36092996;rs4905449;rs10140368;rs8022837;rs34845487;rs35974883;rs12883511;rs2369542;rs8008696;rs8008421;rs1959041;rs34985854;rs11850248;rs11850332;rs11850303;rs11850334;rs55668608;rs35136400;rs12894873;rs11846378;rs11846531;rs11846417;rs11846465;rs11846550;rs71415027;rs71415028;rs2369544;rs28849215;rs28823359;rs12432014;rs61985296;rs12881275;rs7492727;rs34393530;rs35526305;rs34870532;rs72704824;rs112558727;rs66680728 | ENSG00000182512 |
| 14:96600001-96640000 | 14:96000001-96040000 | 1.40E-52 | hESC | rs59804216;rs2369539;rs4627266;rs2369541;rs4905447;rs7156430;rs1889374;rs1889372;rs11160314;rs3939400;rs55940712;rs60634508;rs36024935;rs56334881;rs2151767;rs7144843;rs34485356;rs12888576;rs72704813;rs12894970;rs369235957;rs12885218;rs71415026;rs68023675;rs35262593;rs34857325;rs36092996;rs4905449;rs10140368;rs8022837;rs34845487;rs35974883;rs12883511;rs2369542;rs8008696;rs8008421;rs1959041;rs34985854;rs11850248;rs11850332;rs11850303;rs11850334;rs55668608;rs35136400;rs12894873;rs11846378;rs11846531;rs11846417;rs11846465;rs11846550;rs71415027;rs71415028;rs2369544;rs28849215;rs28823359;rs12432014;rs61985296;rs12881275;rs7492727;rs34393530;rs35526305;rs34870532;rs72704824;rs112558727;rs66680728 | ENSG00000182512 |
| 14:96600001-96640000 | 14:96160001-96200000 | 2.07E-07 | hESC | rs59804216;rs2369539;rs4627266;rs2369541;rs4905447;rs7156430;rs1889374;rs1889372;rs11160314;rs3939400;rs55940712;rs60634508;rs36024935;rs56334881;rs2151767;rs7144843;rs34485356;rs12888576;rs72704813;rs12894970;rs369235957;rs12885218;rs71415026;rs68023675;rs35262593;rs34857325;rs36092996;rs4905449;rs10140368;rs8022837;rs34845487;rs35974883;rs12883511;rs2369542;rs8008696;rs8008421;rs1959041;rs34985854;rs11850248;rs11850332;rs11850303;rs11850334;rs55668608;rs35136400;rs12894873;rs11846378;rs11846531;rs11846417;rs11846465;rs11846550;rs71415027;rs71415028;rs2369544;rs28849215;rs28823359;rs12432014;rs61985296;rs12881275;rs7492727;rs34393530;rs35526305;rs34870532;rs72704824;rs112558727;rs66680728 | ENSG00000100721 |
| Interaction regions (Chromosome:Start base - end base) include one region overlapping one or more GWS SNP in an enhancer and and one region overlapping a gene promoter region. FDR = false discovery rate. | | | | | |

| **S14 Table.** MAGMA gene-based association analysis | | | | | | | | |
| --- | --- | --- | --- | --- | --- | --- | --- | --- |
| **Ensembl Gene ID** | **Gene Symbol** | **Chromosome** | **Gene start position hg19** | **Gene stop position hg19** | **Number of SNPs** | **N** | **Z score** | ***P*-value** |
| ENSG00000124762 | RP11-404P21.8 | 14 | 96621181 | 96780266 | 576 | 49357 | 4.9129 | 4.49E-07 |
| ENSG00000168398 | BDKRB2 | 14 | 96621016 | 96760666 | 528 | 49345 | 4.674 | 1.48E-06 |
| Gene-level association analysis was performed using MAGMA (within FUMA) with the SNP-wide mean model and 1000 Genomes phase 3 release as the reference panel. We considered a window size of ± 50 kb. Genes with p-value < 2.6e-6 (0.05/19,294 gene tests) were considered significantly associated with angioedema. Number of SNPs - the number of SNPs in the input dataset that are annotated to that gene; Z score: the Z-value is a measure of the level of gene association, based on its (permutation) *P* value; *P*-value : the gene association *P*-value, using asymptotic sampling distribution. | | | | | | | | |

| **S15 Table.** Definition of comorbidity from ICD-codes. | |
| --- | --- |
| **Condition** | **ICD-10 code** |
| Hypertension | I109, I15 |
| Diabetes | E10, E11 |
| Congestive heart failure | I50 |
| Myocardial infarction | I21 |
| Chronic renal failure | N18, N19 |
